# Co-development of an evidence-based personalised smoking cessation intervention for use in a lung cancer screening context

**DOI:** 10.1101/2022.07.18.22277751

**Authors:** Harriet D. Quinn-Scoggins, Rachael L. Murray, Samantha L. Quaife, Pamela Smith, Kate E. Brain, Matthew E.J. Callister, David R. Baldwin, John Britton, Philip A.J. Crosbie, Rebecca Thorley, Grace M. McCutchan

**Affiliations:** Division of Population Medicine, School of Medicine, Cardiff University, Cardiff, UK; Academic Unit of Lifespan and Population Health, School of Medicine, University of Nottingham, Nottingham, UK; Wolfson Institute of Population Health, Queen Mary University of London, London, UK; Department of Respiratory Medicine, Leeds Teaching Hospitals NHS Trust, St James’s University Hospital, Leeds, UK; Department of Respiratory Medicine, Nottingham University Hospital, Nottingham, UK; Division of Infection, Immunity and Respiratory Medicine, Faculty of Biology, Medicine and Health, University of Manchester, Manchester, UK

**Keywords:** Lung Cancer, lung cancer screening, emphysema, smoking cessation, intervention, Imaging/CT MRI, patient and public involvement, co-development

## Abstract

**Background:** Optimising smoking cessation services within a low radiation-dose computed tomography (LDCT) lung cancer screening programme has the potential to improve cost-effectiveness and overall efficacy of the programme. However, evidence on the optimal design and integration of cessation services is limited. We co-developed a personalised cessation and relapse prevention intervention incorporating clinical and medical imaging collected during lung cancer screening. The intervention is designed to initiate and support quit attempts among smokers attending screening as part of the Yorkshire Enhanced Stop Smoking study (YESS: ISRCTN63825779). Patients and public were involved (PPI) in the development of an acceptable intervention designed to meet the needs of the target population.

**Methods:** An iterative co-development approach was used. Eight members of the public with a history of smoking completed an online survey to inform the visual presentation of risk information in subsequent focus groups for acceptability testing. Three focus groups (n=13) were conducted in deprived areas of Yorkshire and South Wales with members of the public who were current smokers or recent quitters (within the last year). Exemplar images of the heart and lungs acquired by LDCT, absolute and relative lung cancer risk, and lung age were shown. Data were analysed thematically, and discussed in stakeholder workshops. Draft versions of the intervention were developed, underpinned by the Extended Parallel Processing Model to increase self-efficacy and response-efficacy. The intervention was further refined in a second stakeholder workshop with a PPI panel.

**Results:** Individual LDCT scan images of the lungs and heart, in conjunction with artistic impressions to facilitate interpretation, were considered by public participants to be most impactful in prompting cessation. Public participants thought it important to have a trained practitioner guiding them through the intervention and emphasising the short-term benefits of quitting. Presentation of absolute and relative risk of lung cancer and lung age were considered highly demotivating due to reinforcement of fatalistic beliefs.

**Conclusion:** An acceptable personalised intervention booklet utilising LDCT scan images has been developed for delivery by a trained smoking cessation practitioner. Our findings highlight the benefit of co-development during intervention development and the need for further evaluation of effectiveness.

**PLAIN ENGLISH SUMMARY:** Supporting patients to stop smoking when they attend lung cancer screening will improve the overall benefit and value for money of the service. This study developed a booklet containing pictures of a person’s own lungs and heart taken during a lung cancer screening scan. The booklet shows areas of damage to the heart and lungs caused by smoking, delivered alongside positive messages to build confidence to stop smoking and let patients know about the benefits of stopping smoking.

To develop the booklet, we worked with members of public who currently or used to smoke. Eight members of public completed a survey asking about the best ways to present information about risk. Thirteen members of the public took part in focus groups to co-develop the booklet. One workshop with academic and healthcare professionals and one workshop with a public involvement panel were held to develop and finalise the booklet.

Members of the public said they wanted information about the short-term benefits of quitting smoking, and that coloured drawings next to the scan picture would help them to understand what the scan picture meant. Having someone specially trained to guide them through the booklet was considered important. Being told about their risk for lung cancer in the future was off-putting and might discourage a quit attempt.

We have co-developed a booklet to support people to quit smoking when they go for lung cancer screening. The booklet is currently being tested to see whether it can support people to quit smoking.

## BACKGROUND

Lung cancer has the highest mortality of all cancers in the UK.[1] More than 85% of lung cancer cases are caused by tobacco smoking,[2] and smoking cessation at any age significantly reduces lung cancer risk.[3,4] Lung cancer screening with annual low-dose computed tomography (LDCT) has been shown to reduce lung cancer mortality in high-risk groups by 20%, and is widely available in some high-income countries such as the United States[5]. Results from the Dutch-Belgian NELSON study report substantially lower lung cancer mortality when compared to no screening, even when screening intervals are increased over time.[6] In the UK it is available as part of the Targeted Lung Health Check programme and is currently awaiting a final decision on approval from the UK National Screening Committee.

Empirical evidence suggests that lung cancer screening can act as a ‘teachable moment’ to encourage smoking cessation.[7] Integrating screening and smoking cessation services can improve the overall success and cost-effectiveness of lung cancer screening.[8-13] The UK Lung Screening Trial reported net cessation rates of 15% at two-year follow-up, compared to 4% in the general population.[14] Where smoking cessation interventions have previously been included in lung cancer screening, they have typically been brief and low intensity involving signposting to external services (e.g. 15). Low intensity interventions are likely to be inadequate for the population of long-term (and often life-long) smokers eligible for lung cancer screening who typically have higher dependence on nicotine,[16] less success in quitting,[17] and are over-represented in socioeconomically deprived groups where cessation is most challenging.[18] Novel, higher intensity smoking cessation interventions may be needed to support and sustain quit attempts in individuals at high risk of developing lung cancer; such interventions delivered at the point of screening have the potential to increase cessation rates in socioeconomically deprived populations [19-21]. Interventions should be developed in consultation with eligible members of the public to ensure the intervention developed meets the needs of the target population [22].

Research outside of the lung screening setting suggests that interventions using visual or imaging techniques can effectively promote smoking cessation.[23,24] Visual feedback has the potential to strengthen risk communication as imaging results can reveal visible evidence of bodily harm attributable to smoking that is immediately comprehensible to the patient.[24,25] Using clinical information and imaging obtained from LDCT scans during a lung cancer screening appointment could therefore prove an effective motivational tool, providing a higher intensity, personalised intervention better suited to the high-risk target population, especially when developed in collaboration with potential service users. The current study was underpinned by the Extended Parallel Process Model (EPPM; [26]) as a conceptual framework for understanding individual variation in the behavioural response to receiving personal risk information. The EPPM proposes that when a high level of threat is perceived, an individual will only engage in protective behaviours (‘danger control’) if they are confident in their ability to enact the behaviour (self-efficacy) and they believe that the behaviour will be effective in reducing their risk (response efficacy). In a scenario where these efficacy beliefs are low, an individual will respond defensively to high threat information and engage in avoidant behaviours that reduce their negative emotional arousal (‘fear control’). In the context of lung cancer screening, personalised risk interventions should therefore be designed to enhance perceptions of self-efficacy and response-efficacy if they are to effectively motivate smoking cessation efforts in response to the threat associated with visualising personal risk of heart and lung disease.

The current study is the first to report the co-development of a bespoke smoking cessation and relapse prevention intervention for the Yorkshire Enhanced Stop Smoking Study (YESS; ISRCTN63825779 [27]), underpinned by the EPPM [26] and embedded within the Yorkshire Lung Screening Trial (YLST; ISRCTN42704678 [28]). YLST aims to describe participation among people at highest risk of lung cancer and clarify the optimal strategy for defining a high-risk population for screening in the. YESS is currently testing the effectiveness of the intervention reported and how to best integrate smoking cessation in a lung cancer screening setting.

Following the principles of co-production in healthcare and the MRC complex intervention development guidance,[22,29] key stakeholders were involved (members of the public, health professionals and academic partners) to co-develop the materials for the YESS intervention - a personalised risk information booklet (including images of the participant’s scan from their screening appointment alongside personalised risk text and practitioner scripts; both designed to enhance self-efficacy and response-efficacy) and personalised on-going support provided by a trained smoking cessation practitioner. Members of the public were involved throughout intervention development to ensure that the booklet was acceptable to the target population to potentially increase overall impact and effectiveness of the intervention.

## METHODS

Intervention materials were developed iteratively. Acceptability and format preferences regarding personalised risk intervention components (absolute and relative lung cancer risk, lung scan images showing emphysema, heart scan images showing coronary heart calcification, and lung age) were collected sequentially through a series of focus groups. Prior to this, the selection of images to be presented in the focus groups was initially informed by the findings of an online survey involving members of the public who smoke. Intervention materials were further refined through two stakeholder workshops. Participants were shown hypothetical examples of risk information and advised that the final intervention would include images based on their individual clinical data from lung cancer screening. Examples of visual risk materials presented by intervention component are provided in Figure 1. All visual risk materials, survey questions and focus group topic guides are provided in Additional Files 1 and 2. Ethical approval for the study was gained from the Cardiff University School of Medicine Research Ethics Committee (ref 17/51).

**Figure 1.**
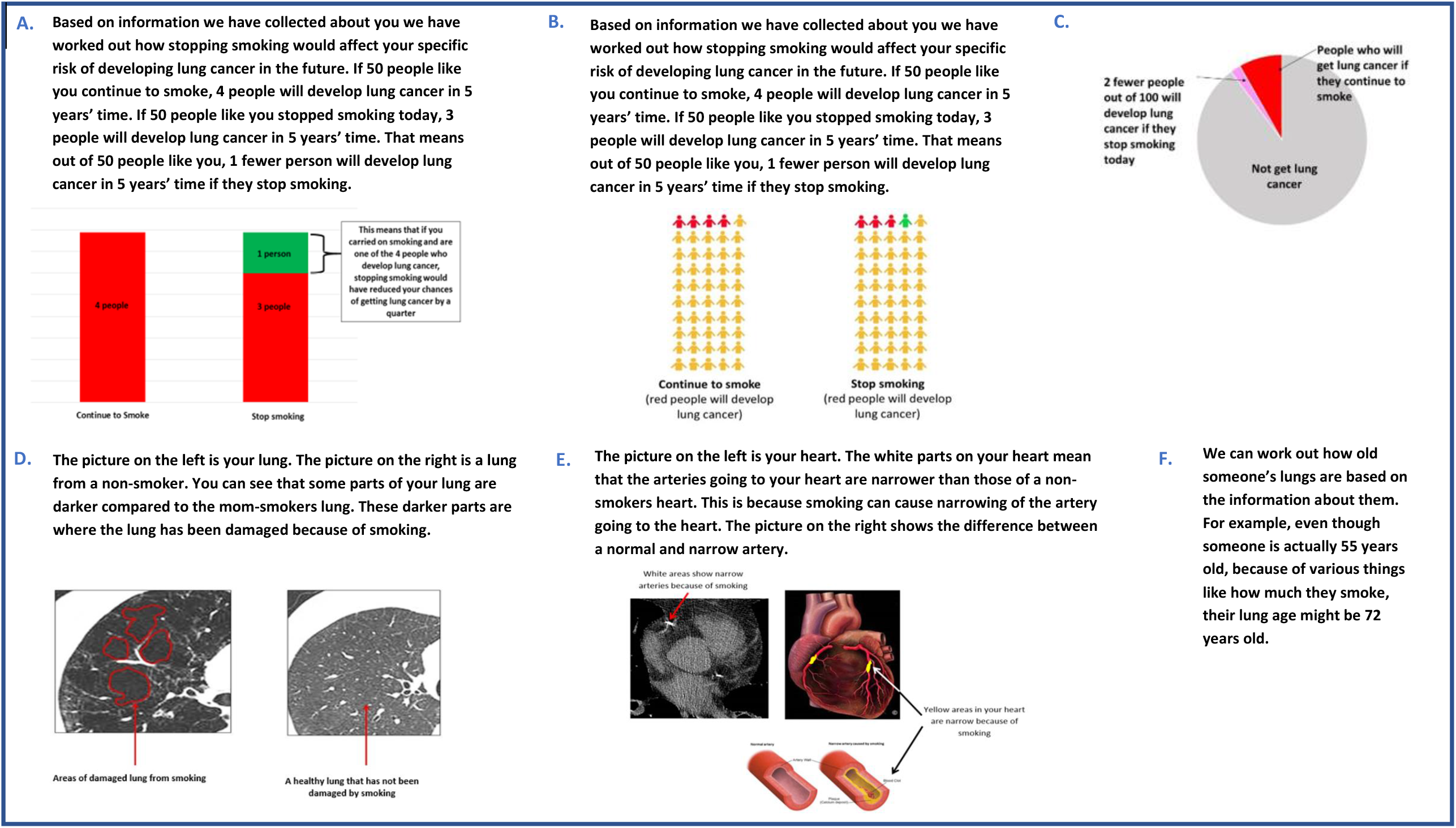
Examples of Online Survey and Focus Group Materials **A**. Bar chart showing absolute and relative lung cancer risk **B**. Pictogram showing absolute lung cancer risk **C**. Pie chart showing absolute lung cancer risk **D**. Lung scan images showing emphysema **E**. Heart scan images showing coronary artery calcification **F**. Lung age text

### Online Survey

An online survey was conducted to inform the selection of visual presentations of lung cancer risk that were subsequently presented for feedback in the focus groups. The online survey (Additional File 1) was distributed via Survey Monkey to an established patient and public involvement group for tobacco related research in the East Midlands, convened by the UK Centre for Tobacco and Alcohol Studies. Survey participants were asked to select their preferred visual representation of absolute risk and relative risk for lung cancer (pie chart, bar chart or pictogram), framing (gain, loss or partial framing), time frame to present absolute lung cancer risk reduction (2 years, 5 years, 7 years or 10 years), and pictogram format (number and colouring of icons). A free text box and researcher contact information were provided for gathering additional comments.

### Focus Groups

Three focus groups (two in South Wales and one in Yorkshire) were conducted with people over age 55 years who currently smoked or had quit within the past year, recruited from highly deprived areas (assessed by multiple deprivation score) through pre-existing community contacts and snowball sampling.[30] Individual level deprivation (educational attainment) was assessed during data collection. Preferred visual presentations of risk information from the online survey informed topic guide development (Additional File 2). Members of the public were asked to provide feedback on format preferences regarding: (1) personalised scan images of lungs with emphysema presented alongside artist impressions of healthy areas of the lung, and as axial/dorsal views; (2) personalised scan images of the heart showing coronary artery calcification presented alongside a healthy heart, artist impressions of the heart, and a 3D representative heart with coloured sections to highlight calcification; (3) absolute and relative risk for lung cancer shown as bar charts or pictograms, and (4) lung age shown as a textual description. Focus groups were audio-recorded and transcribed verbatim. Thematic analysis [31] was conducted using NVivo 11 Qualitative Data Analysis Software.

### Stakeholder Workshops

The final intervention was developed and refined in accordance with findings from the focus groups, existing literature and in two workshop-style meetings with stakeholders. The first stakeholder workshop included representation from health psychologists, clinicians, specialist research nurses, specialists in health policy, and researchers with extensive knowledge and experience working in cancer screening and smoking cessation. Each of the intervention components were presented to stakeholders in turn, showing an exemplar of how the information could be presented visually, alongside existing evidence for the component and findings from the focus groups with members of the public. In a facilitated discussion, stakeholders were asked to comment on the suitability of each intervention component in the YESS context and acceptability/feasibility of intervention delivery. The second stakeholder workshop was conducted with public and patient representatives from the Nottingham Smokers Panel. A draft version of the booklet was shown to the Nottingham Smokers Panel for feedback in a facilitated discussion.

### Patient and Public Involvement

Members of the public were involved throughout intervention development, in accordance with a co-development approach. A co-development approach was utilised to ensure that the intervention was developed to meet the needs of the target population. The near-final version of the intervention was presented to a patient and public involvement panel of people with a history of smoking. The Guidance for Reporting Involvement of Patients and Public checklist guided reporting (Additional File 3; [32]).

## RESULTS

### Online Survey

Eight participants completed the online survey. Pictograms were favoured by five participants over the bar chart (n=2) and pie chart (n=1) for visualisation of risk. There was no clear preference for the time frame that would be most meaningful to present risk information (three years n=2, five years n=2, seven years n=2, ten years n=2). Pictograms that presented lung cancer risk in icon arrays of 100 were favoured over icon arrays of 50 by six participants, with positive effects on lung cancer risk with smoking cessation shown in the pictogram as green (n=5). Six participants preferred gain framed statements, whereas two preferred partial framing.

### Focus Groups

Thirteen members of the public participated across three focus groups, of whom nine currently smoked and four had recently quit. Results are presented according to each intervention component, with exemplary quotes.

#### Lung cancer absolute and relative risk

Members of the public who took part in the focus groups described absolute risk information for lung cancer as demotivating because it undermined the perceived benefit of quitting. The risk was not considered high enough to make a difference to their smoking behaviour or prompt a quit attempt. Although relative risk information was understood with ease, they focused on the seemingly small absolute risk benefit of smoking cessation. How lifetime risk could be calculated was queried, and most were concerned about being told their life expectancy.

> *‘… You go through all that giving up and it’s only one in four…’* (Participant 3, Focus Group 1)

> *‘… I think looking at those odds, that wouldn’t bother me, I’d carry on… It’s not the fear factor there…’* (Participant 4, Focus Group 3)

> *‘… I don’t think this chart frightens you enough, it’s just a chart. You have to put people here like they are real…’* (Participant 6, Focus Group 1)

Fatalism was described with reference to knowing their lung cancer risk was high, and participants suggested that this would evoke a fearful avoidant response instead of encouraging a quit attempt.

> *‘…Well if I tried to stop smoking and I couldn’t I’d probably say c’est la vie and carry on smoking and it wouldn’t. I thought of dying as a result of smoking and the slow death that will be endured if the lungs go and other parts of your body go and then you get cancer and all that sort of, and whatever else…’* (Participant 3, Focus Group 2)

##### Lung LDCT scan images showing emphysema

Lung LDCT scan images were well received. A scan image of their lungs with coloured highlighted areas of damage presented alongside a library image of a healthy lung for comparison was preferred. The scan image was difficult to interpret alone; participants highlighted the importance of accompanying text and verbal explanation by a trained advisor.

> *‘…I think the difference between these two is the one on the right looks like a lung. The one of the left just looks like a black and white picture. It could be a picture of any part of your body unless you were a specialist you wouldn’t know if you take the writing away…’* (Participant 2, Focus Group 2)

> *‘… What we want is an internal picture, coloured picture of his lung and then mine to compare…’* (Participant 2, Focus Group 1)

There was a preference for scan images of lungs to bar charts depicting risk, and some suggested that scan images were a better way of communicating risk. Scan images were considered potentially motivating for cessation because the images would be their own, meaning they were less able to ignore the information or deny personal relevance.

> *‘… But the lung thing is genuine, that’s what you’re breathing in. If you’re inhaling smoke and breathing in, that is causing a problem. So a similar sort of picture but of the lung would be definitely a lot better…’* (Participant 2, Focus Group 1)

> *‘… Yeah because at the end of the day we’re smokers we’re a bunch of pessimists. We see someone else’s lung, that’s not mine. If there was a picture of your own lung, you can’t deny the fact I think…’* (Participant 1, Focus Group 30

##### Heart LDCT scan images showing coronary artery calcification

The 3D coloured image of the heart highlighting calcification was preferred over black and white scan images. Small sections showing coronary artery calcification on the 3D heart scan image were easier to interpret when shown alongside an artist’s impression of a healthy and calcified artery.

> *‘…The two together are good because the picture of the whole heart, the narrow parts are very small and highlighted by the white. But with the expanded view of the two that gives much more detail…’* (Participant 2, Focus Group 2)

##### Lung age

There was good comprehension that smoking increases lung age, and that increased lung age may suggest early mortality. However, the impact of lung age on motivation to quit was viewed negatively due to interpretation of lung damage as irreversible, evoking a fearful emotional response. Most reported that lung age data would not motivate them to quit smoking.

> *‘… It races your life too far into the future when you’re only 55. It advances you closer to death…’* (Participant 3, Focus Group 2)

> *‘… You can’t make it better can you… it’s irreversible…’* (Participant 3, Focus Group 1)

> *‘… It’s frightening to see that but I don’t think that would stop me smoking…’* (Participant 1, Focus Group 2)

#### Stakeholder Workshops

Intervention refinements were made through stakeholder feedback. In the first stakeholder workshop with academics and health professionals it was agreed that personalised lung and heart scan images alongside explanatory artistic impressions, delivered with the support of a trained advisor alongside standardised scripted verbal advice to boost self- and response-efficacy, would be included in the YESS intervention. Absolute and relative lung cancer risk and lung age were removed to mitigate against any potential negative effects in accordance with advice from members of the public who took part in the focus groups. Following feedback from the second stakeholder workshop a cost calculator to show how much money would be saved by quitting was removed because patient and public involvement smoking panel representatives considered it to be naive and suggested it was likely to make patients defensive and dismissive. Additionally, minor presentation changes were made to the booklet in accordance with suggestions from the patient and public panel representatives (e.g. images of exemplary patients were removed from the front cover). Presentation adjustments were made to aid readability and comprehension, and to increase personalisation.

## DISCUSSION

To our knowledge, this study is the first to report the co-development of a bespoke visual smoking cessation and relapse prevention intervention for integration in targeted lung cancer screening. Our findings indicate that in high-risk adults living in socioeconomically deprived areas, personalised risk interventions using LDCT scan images, alongside scripted verbal explanation that targets efficacy perceptions and is delivered by trained smoking cessation practitioners (Table 1), could motivate smoking cessation in a lung cancer screening context. By involving patients and members of the public throughout intervention development and using a co-development approach, we have developed an acceptable personalised smoking cessation intervention for use in a lung cancer screening setting (Additional File 4).

**Table 1.**
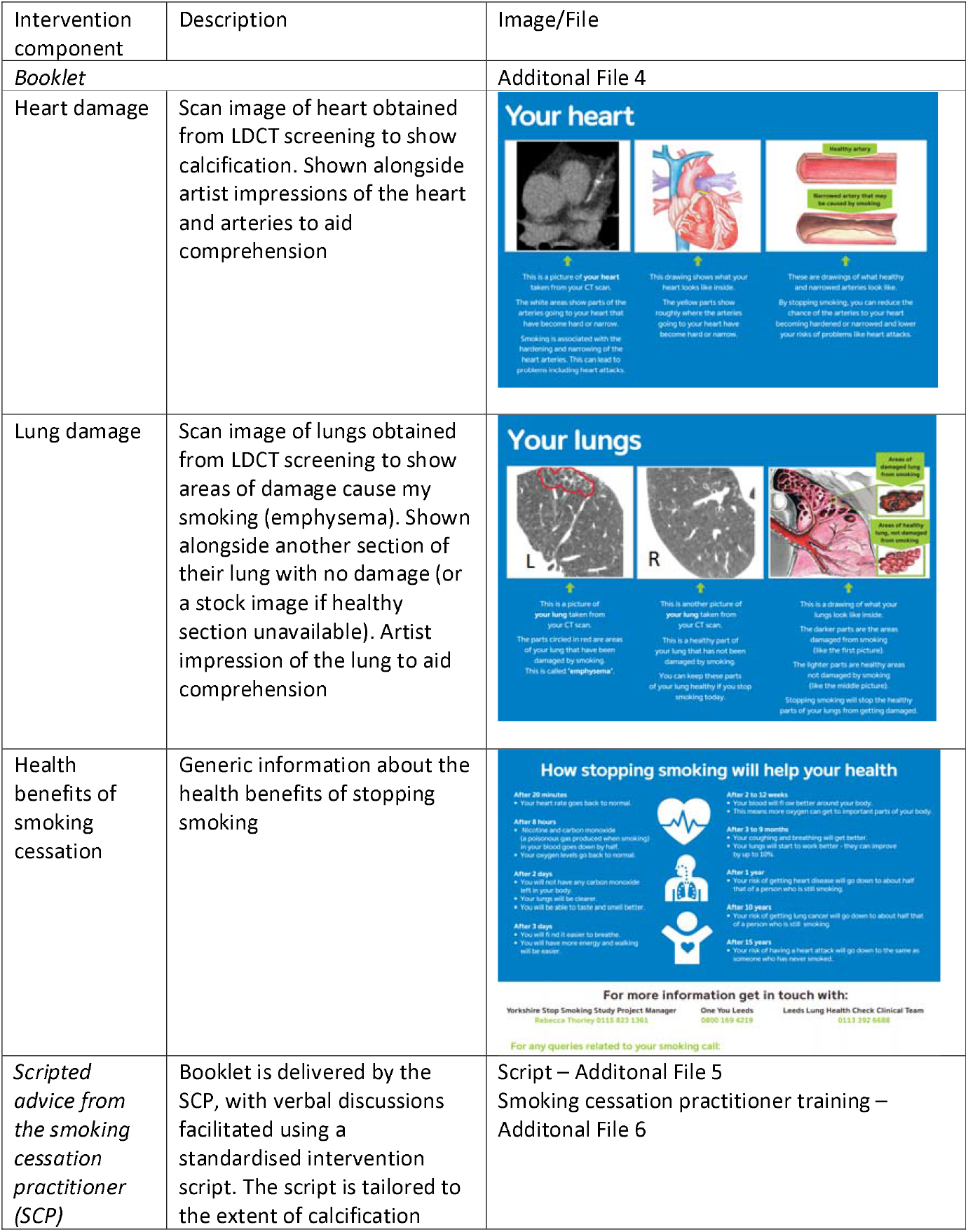
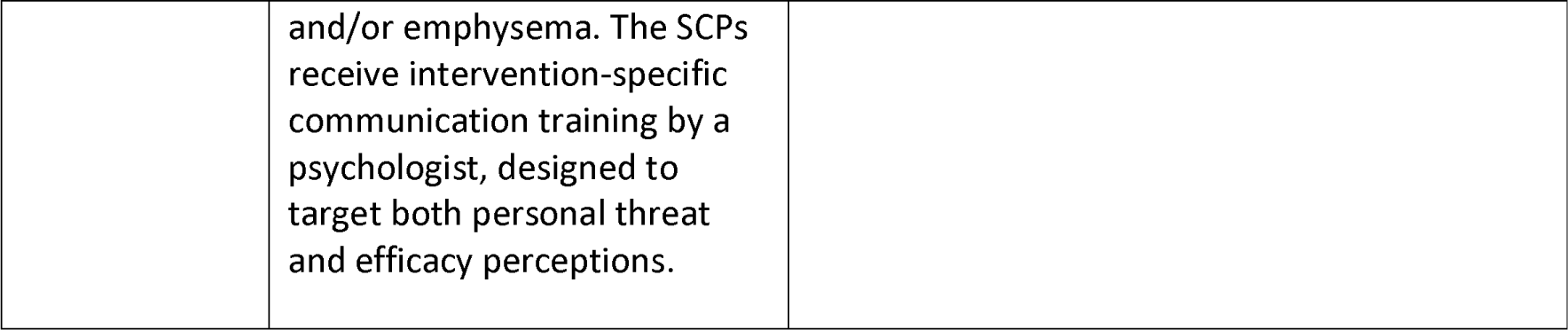
YESS intervention components.

Through a three-pronged intervention co-development approach (identifying the evidence base, patient and pubic involvement, and stakeholder feedback), we have developed an intervention that meets the needs of the target population. Patient and public involvement is especially important when considering the sensitive and unique nature of the intervention and context of delivery in this case. Not only are this high-risk population taking part in lung cancer screening, but with their likely higher dependence on nicotine [17] and lower success in quitting,[18] it is essential that fatalistic beliefs or blame are not reinforced by the intervention. We incorporated the views and preferences of the target population to increase acceptability, relevance and engagement of the final intervention.

Results suggest that scan images were perceived as relevant and easy to understand, and were considered most likely to motivate a quit attempt because recipients would be unlikely to disregard risk information based on visual imaging of their own lungs and heart. Although empirical evidence regarding the effectiveness of medical images to promote health behaviour change is mixed,[33] interventions using medical images have been shown to be effective when combined with physician’s advice [24]. A systematic review found that presenting projected health risk estimates alone, even when highly personalised, did not produce strong effects on health-related behaviours or sustained change.[21] Similarly, a Cochrane Review assessed the effectiveness of biomedical risk assessments as an aid for smoking cessation (including biofeedback on risk exposure, smoking-related disease and smoking-related harm) and found no evidence of increased cessation rates;[34]. This highlights the importance of providing on-going behavioural support from a trained smoking cessation practitioner (Additional File 5).

According to the EPPM, the way in which medical images are presented and the content of the accompanying advice are important in determining the behavioural impact of personalised medical images. The intervention needs to not only be effective in communicating personal salience of risk, but also in building self-efficacy and response-efficacy. The intervention should then sufficiently increase threat perceptions in a way that motivates individuals to engage in risk-reducing behaviour because they also feel able to quit and that quitting will improve their health. The YESS intervention includes communication techniques and language designed to target both personal threat and efficacy perceptions, interwoven throughout the intervention booklet and verbal discussions with the trained smoking cessation practitioner. Verbal discussions are facilitated using a standardised intervention script (Additional File 5) delivered by the smoking cessation practitioner, who is also given intervention-specific communication training by a psychologist (Additional File 6). The scripts and discussions are tailored to the extent of calcification and/or emphysema.

Members of the public found pictorial representations of absolute/relative risk and lung age harder to understand and considered them demotivating for encouraging a quit attempt because they reinforced fatalistic beliefs that lung damage was irreversible, undermining the perceived response efficacy of smoking cessation. Previous research outcomes on the presentation of personalised risk messages and projected long-term risk of health conditions in smoking cessation for current smokers is varied when compared to provision of standardised general information.[35-37] Results from these studies suggest that the impact of risk perceptions could be moderated by risk status placed on smoking and smoking products by wider society.[35] It also suggested that personalised risk messages can increase engagement in stop smoking services and short-term abstinence; however, no evidence currently exists about how this translates to sustained quit rates.[36,37]

The current findings suggest that presenting lung age to the target group for lung cancer screening may have unintended adverse consequences in discouraging smoking cessation, in contrast to evidence from previous studies involving younger and more affluent groups. For example, Parkes et al. reported increased likelihood of a quit attempt when lung age was provided to current smokers, however participants in this study were younger (with an average age of 53).[38] It is possible that in a population of older adults experiencing high levels of socioeconomic deprivation, lung age can be difficult to interpret and demotivating when taking into consideration general low expectations of health, life expectancy and well-being.[39]

Our findings reinforce the importance of emphasising the proximal short-term benefits of cessation with the target population to increase response efficacy. Participants highlighted the importance of providing verbal explanations delivered by trained practitioners to increase understanding of the scan images and reinforce the immediate and concrete gains to be made from quitting smoking. Shorter-term future orientation is not only important when providing information on personal lung cancer risk and screening results, but in the delivery of behaviour change techniques to invoke quit attempts and cessation. When taking into account the competing influences and wider social determinants of health [40] that can act as barriers to achieving distal goals, short-term attainable goals with high quality goal setting, are viewed to be more achievable and can thus lead to instant results.[41,42] This short-term orientation may strengthen an individual’s perceptions of both their self-efficacy for reducing risk and the response-efficacy of smoking cessation for their health; perceptions that the EPPM proposes are instrumental to achieving behaviour change in personalised risk interventions.

There were limitations to our study. We were unable to obtain further demographic details from the relatively small sample of online survey participants. Due to difficulties associated with the recruitment of current smokers to a focus group about a smoking cessation intervention, we sampled both people who currently smoke and people who recently quit smoking; however, former smokers included were those who had quit less than one year prior to data collection to ensure that we garnered views from people who had recent experience of smoking cessation.

## CONCLUSION

Smoking cessation interventions that incorporate patients’ scan images to highlight the personalised benefits of cessation, in combination with supportive conversations with a trained practitioner designed to enhance self-efficacy and response-efficacy, could motivate and sustain quit attempts in people who currently smoke participating in lung cancer screening. The personalised cessation and relapse prevention intervention is being evaluated in the YESS study as part of YLST.

## Data Availability

Data sharing is available upon reasonable request. Please contact the corresponding author. The intervention materials are included as additional files.

## LIST OF ABBREVIATIONS

LDCT: low-dose computed tomography
YESS: Yorkshire Yorkshire Enhanced Stop Smoking
YLST: Yorkshire Lung Screening Trial
EPPM: Extended Parallel Processing Model

## DECLARATIONS

### Ethical approval and consent to participate

Ethical approval was provided by Cardiff University School of Medicine Research Ethics Committee (ref 17/51). Written consent was obtained from all participants.

### Consent for publication

Written consent to publish quotes from the qualitative research was obtained from all participants.

### Author contributions

Materials for the online survey and focus groups were drafted by GM. RM, KB, MC and SQ provided substantial input and expert advice to develop and refine draft materials that were shown in the online survey and focus groups. All authors contributed to the development and refinement of intervention materials. GM and PS conducted the focus groups, and PS conducted the focus group data analysis. HQS drafted the manuscript and all authors contributed to the review and editing of the manuscript. All authors read and approved the final manuscript.

## Acknowledgements

The authors gratefully acknowledge all participants who took part in focus groups and completed the online survey. We would like to thank the Nottingham Tobacco and Nicotine Discussion Group for their contribution to the project. Thanks to the YESS trial management group (Alexandra Ashurst, David Baldwin, Kate Brain, John Britton, Matthew Callister, Phillip Crosbie, Nicola Hawkes, Sarah Lewis, Monica Londahl, Grace McCutchan, Rachael Murray, Richard Neal, Steve Parrott, Samantha Quaife, Harriet Quinn-Scoggins, Suzanne Rogerson, Pamela Smith, Rebecca Thorley, Qi Wu) for their continued support with the project.

