## Additional File 1 for "Co-development of an evidence-based personalised smoking cessation intervention for use in a lung cancer screening context"

### Additional File 1: Online Survey

Five examples were manipulated on the survey to explore:

1. Preferences on format (bar chart vs pictogram vs pie chart)

2. Time frame to present risk information (3years vs 5 years vs 7 years vs 10 years)

3. Preferred denominator (out of 50 people vs out of 100 people)

4. Preferences on gain or loss framing

5. Preferences on partial gain framing

A free text box and researcher contact information was also provided for participants to provide any further comments on the above.

#### Examples provided in Online Survey

1. Which option do you prefer for the following information:

   'If 100 people like you continue to smoke, 8 people will develop lung cancer in 5 years' time. If 100 people like you stopped smoking today, 6 people will develop lung cancer in 5 years' time. That means out of 100 people like you, 2 fewer people will develop lung cancer in 5 years' time if they stop smoking'

   Please order the formats from your most to least favourite (1= most favourite, 2=second favourite, 3= least favourite).

   Option 1A: Bar Graph

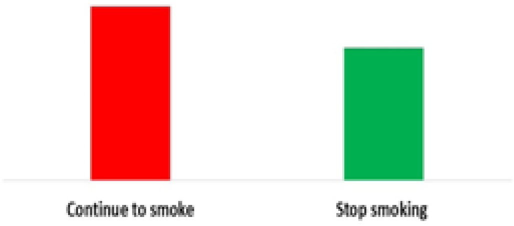


Option 1B: Picture

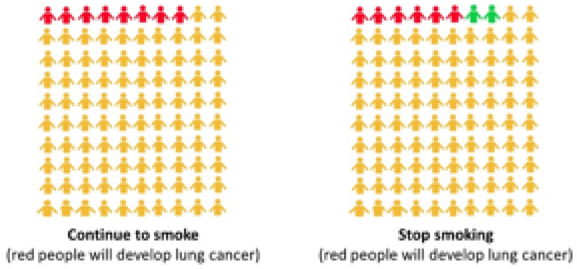


Option 1C: Pie Chart

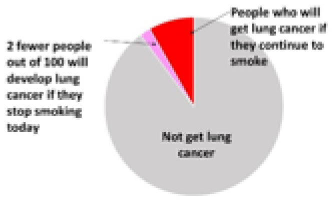

2. How far ahead do you think people would like to know what chances of getting lung cancer? Please order from 1 (people would most like to know this amount of time) to 4 (people would least like to know this amount of time)

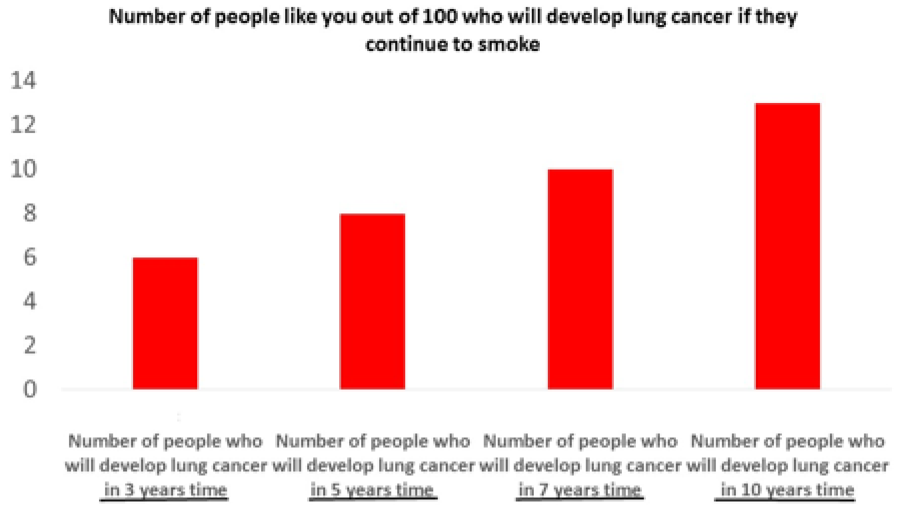

3. Which of the following pictures do you prefer?

Option 3A - Picture with 100 people

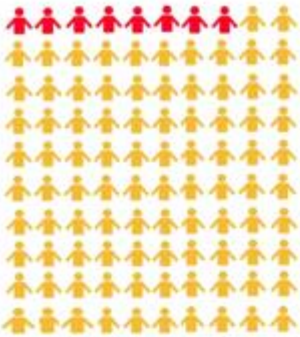

**Continue to smoke**
(red people will develop lung cancer)

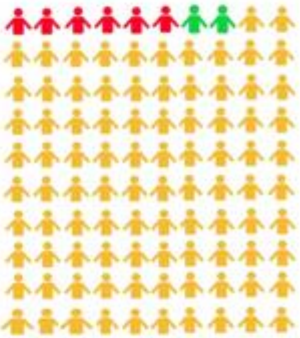

**Stop smoking**
(red people will develop lung cancer)

Option 3B: Picture with 50 people

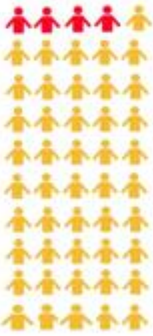

**Continue to smoke**
(red people will develop lung cancer)

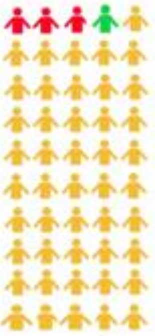

**Stop smoking**
(red people will develop lung cancer)

4. Which statement do you prefer?

**Option 4A- The benefits of quitting smoking**

'If 100 people like you stopped smoking today, 2 fewer people will develop lung cancer in 5 years time compared to if you carried on smoking.'

**Option 4B- The harms of continuing to smoke**

'If 100 people like you carried on smoking, 8 people will develop lung cancer in 5 years time.'

5. Which picture do you prefer?

Option 5A - Pictures of 100 people showing difference in green

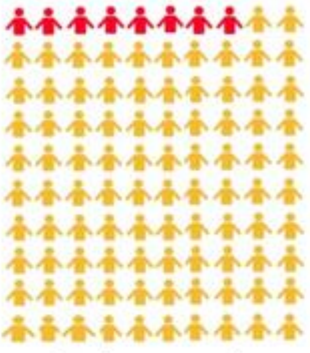

**Continue to smoke**
(red people will develop lung cancer)

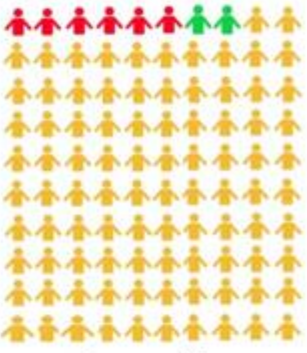

**Stop smoking**
(red people will develop lung cancer)

Option 5B – Pictures of 100 people without showing difference in green

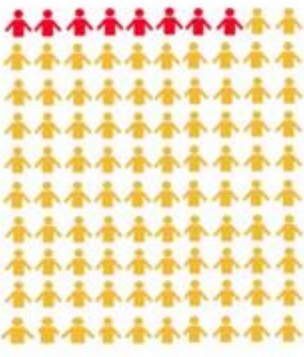

**Continue to smoke**
(red people will develop lung cancer)

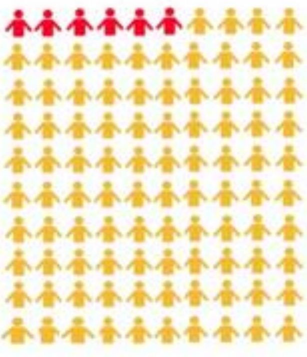

**Stop smoking**
(red people will develop lung cancer)

6. Do you have any other comments or suggestions?

7. Please provide your name and phone number here so that Grace can ring you to discuss your preferences in more detail. If you would prefer not to speak on the phone, just leave this section blank.

8. We will pick someone at random to win a £25 shopping voucher. Please write your name and contact details in the box below if you would like to be entered into the draw for a chance to win a £25 shopping voucher (optional)
