## Additional File 2 for "Co-development of an evidence-based personalised smoking cessation intervention for use in a lung cancer screening context"

### Additional File 2 - Focus Groups

Four intervention components were discussed in the focus groups to explore:

1. Absolute risk and relative risk for lung cancer
    a. Five year absolute risk reduction for lung cancer (continue to smoke vs smoking cessation; shown as a bar chart and pictogram)
    b. Five year absolute and relative risk reduction for lung cancer (continue to smoke vs smoking cessation; shown as a bar chart)
    c. Preferences for time frame to present absolute lung cancer risk reduction (2 years vs 5 years vs 10 years)
2. Lung scan images showing emphysema
    a. Areas of damage due to smoking vs healthy areas of the lung. Format preferences included showing areas of their damaged lung next to a library image of a healthy lung vs healthy areas of their own lung.
    b. Areas of damage due to smoking next to an artist's impression showing healthy and damaged areas of the lung
    c. Image of lung: axial vs coronal image
3. Heart scan images showing coronary artery calcification
    a. Coronary artery calcification due to smoking vs a library image of a healthy heart with no coronary artery calcification. Format preferences included showing scan images of their heart vs a representative image of their heart
    b. Areas of coronary artery calcification damage due to smoking next to an artist's impression of magnified arteries showing a healthy artery and calcified artery due to smoking
    c. Three image options for preferred representative image of the heart

Lung age

#### Examples provided in Focus Group 1

##### Example 1a

'The picture on the left is your lung. The picture on the right is a lung from a non-smoker. You can see that some parts of your lung are darker compared to the non-smokers lung. These darker parts are where the lung has been damaged because of smoking.'

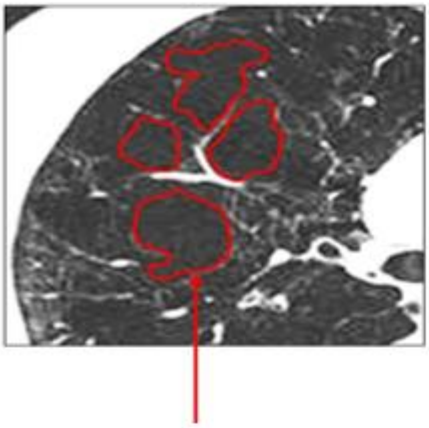

Areas of damaged lung from smoking

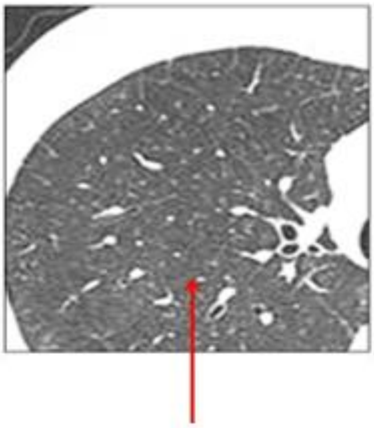

A healthy lung that has not been damaged by smoking

##### Example 1b

'Both of these pictures are your lungs. The darker parts on the picture on the left show the damage in your lungs because of smoking. The picture on the right shows healthy parts of your lung. These healthy parts have not yet been damaged by smoking.'

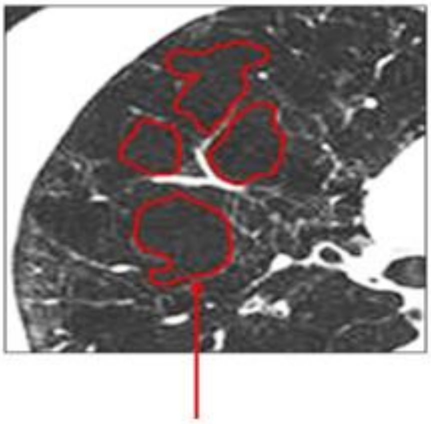

Areas of damaged lung from smoking

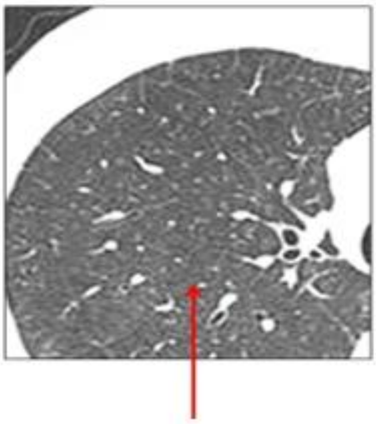

Healthy parts of your lung that have not been damaged from smoking

#### Example 2

'The picture on the left is your lung. The darker parts on your lung have been damaged because of smoking. The picture on the right is a drawing of a lung. On this drawing, some parts have been damaged by smoking, and other parts are healthy.'

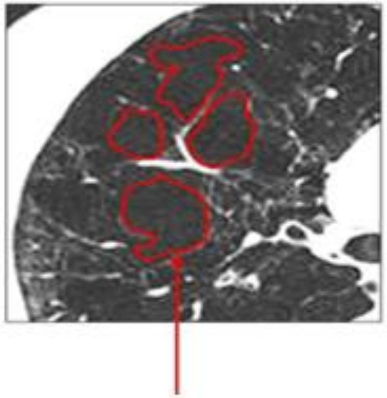

Areas of damaged lung from smoking

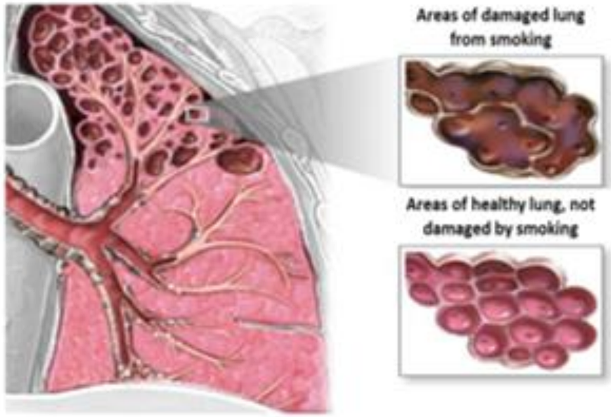

**Example 3a**

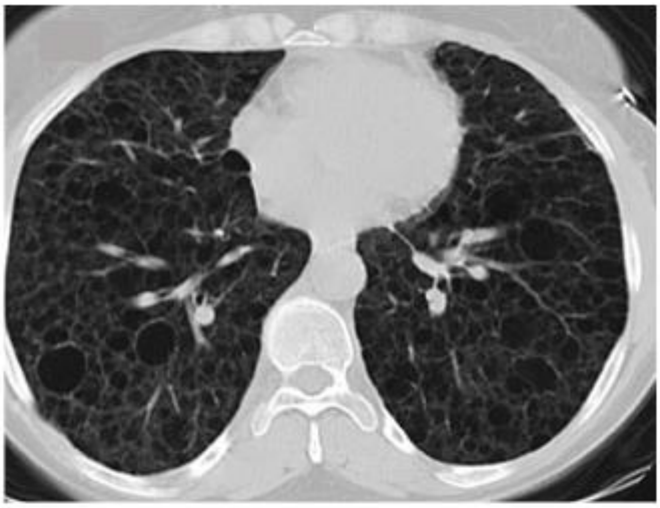

**Example 3b**

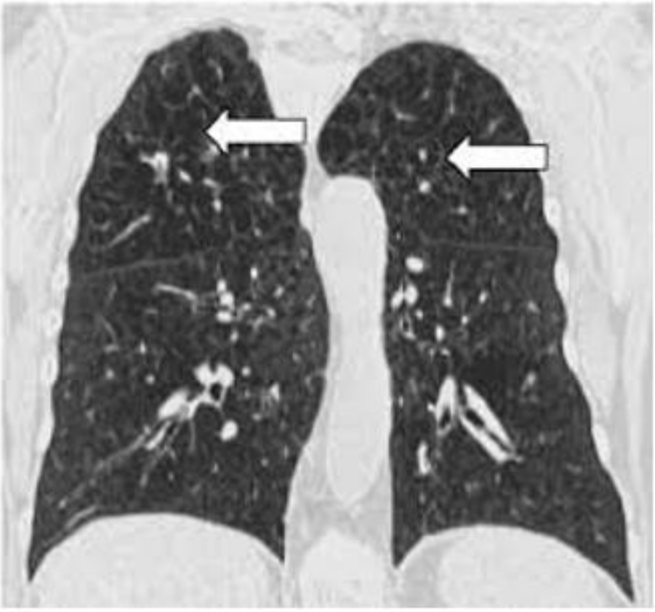

**Example 4**

**‘We can work out how old someone’s lungs are based on the information about them. For example, even though someone is actually 55 years old, because of various things like how much they smoke, their lung age might be 72 years old’**

#### Example 5

**'The picture on the left is your heart. The picture on the right is a heart from a non-smoker. The yellow or white parts on your heart mean that the arrteries going to your heart are more narrow than those of non-smokers heart. This is because smoking can cause narrowing of the artery going to the heart.'**

Your heart

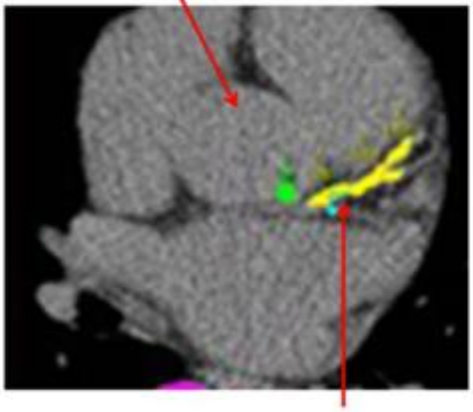

Yellow shows narrow arteries to the heart because of smoking

A normal heart

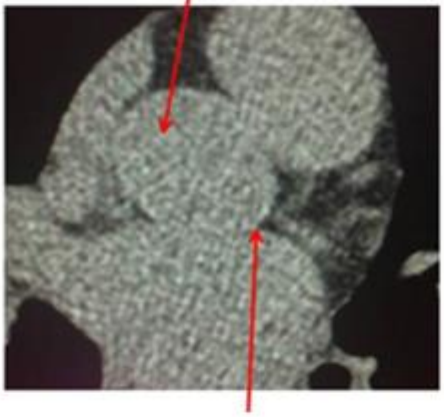

No narrowing of the arteries to the heart

**Representative picture of your heart**

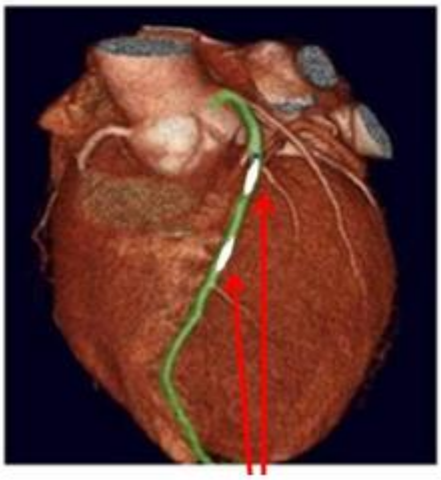

White shows narrow arteries to the heart because of smoking

**A normal heart**

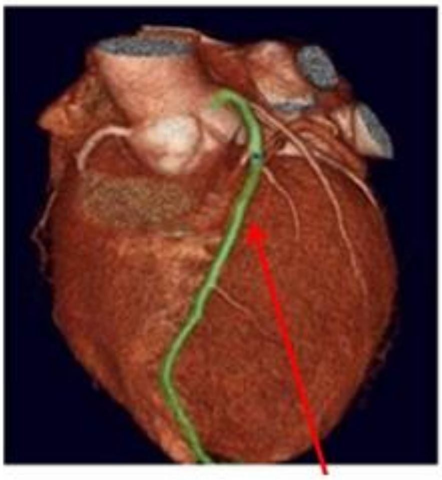

No narrowing of the arteries to the heart

#### Example 6

'The picture on the left is your heart. The white parts on your heart mean that the arteries going to your heart are narrower than those of non-smokers heart. This is because smoking can cause narrowing of the artery going to the heart. The picture on the right shows the difference between a normal and narrow artery.'

Representative picture of your heart

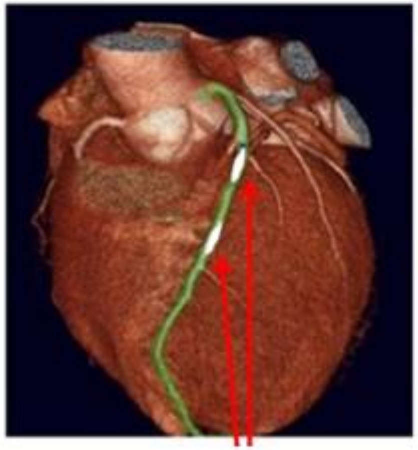

White shows narrow arteries to the heart because of smoking

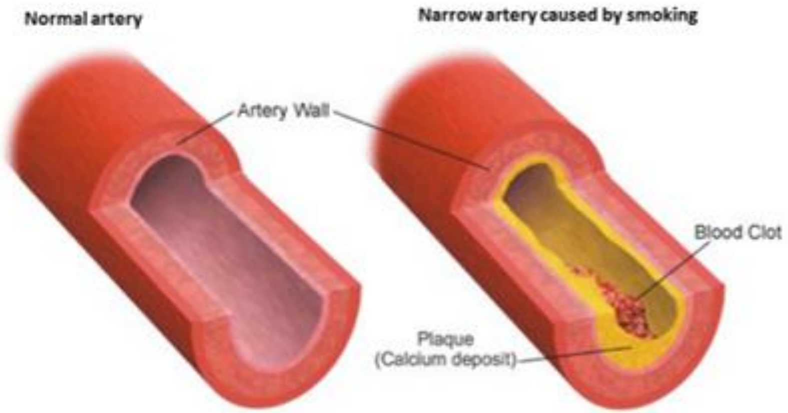

**Example 7a**

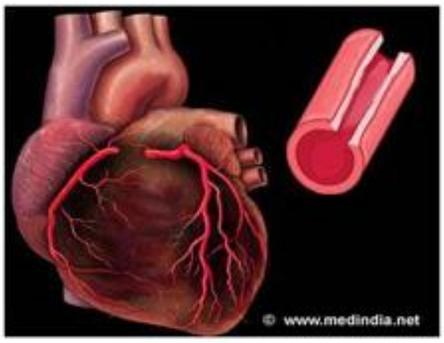


**Example 7b**

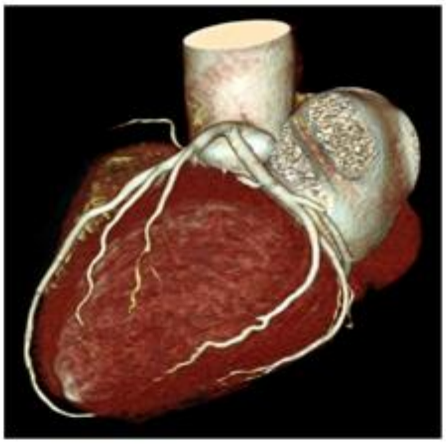

**Example 7c**

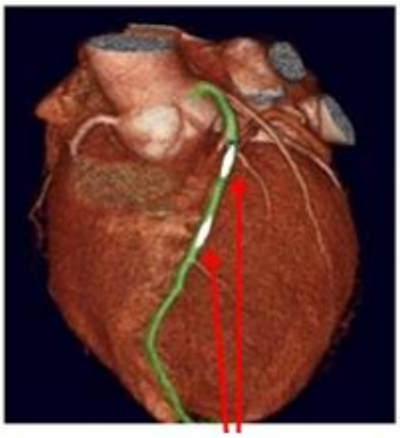

#### Example 8

'Based on information we have collected about you, we have worked out how stopping smoking would affect your specific risk of developing lung cancer in future. If 50 people like you continue to smoke, 4 people will develop lung cancer in 5 years' time. If 50 people like you stopped smoking today, 3 people will develop lung cancer in 5 years' time. That means out of 50 people like you, 1 fewer person will develop lung cancer in 5 years' time if they stop smoking'

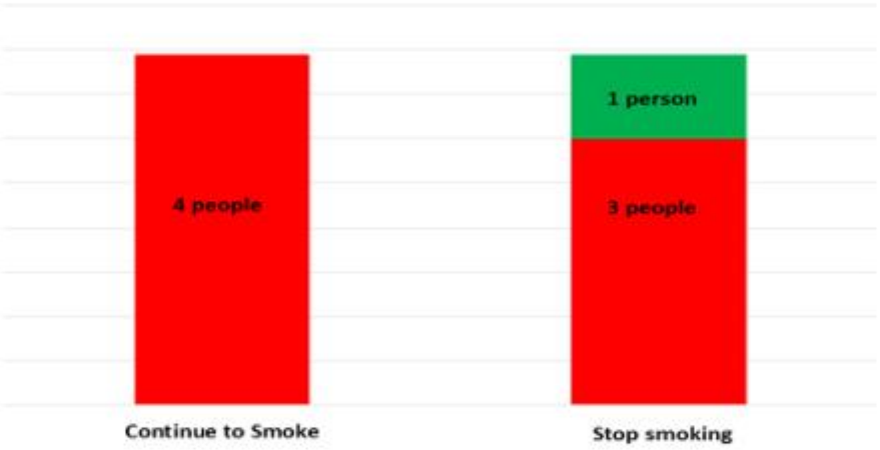


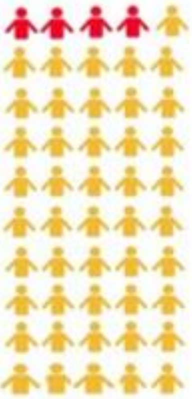

**Continue to smoke**
(red people will develop lung cancer)

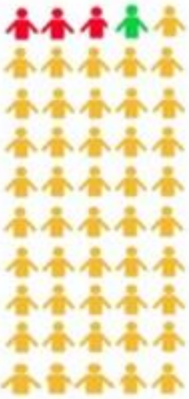

**Stop smoking**
(red people will develop lung cancer)

**Example 9**

**'Based on information we have collected about you, we have worked out how stopping smoking would affect your specific risk of developing lung cancer in future. If 50 people like you continue to smoke, 4 people will develop lung cancer in 5 years' time. If 50 people like you stopped smoking today, 3 people will develop lung cancer in 5 years' time. That means out of 50 people like you, 1 fewer person will develop lung cancer in 5 years' time if they stop smoking'**

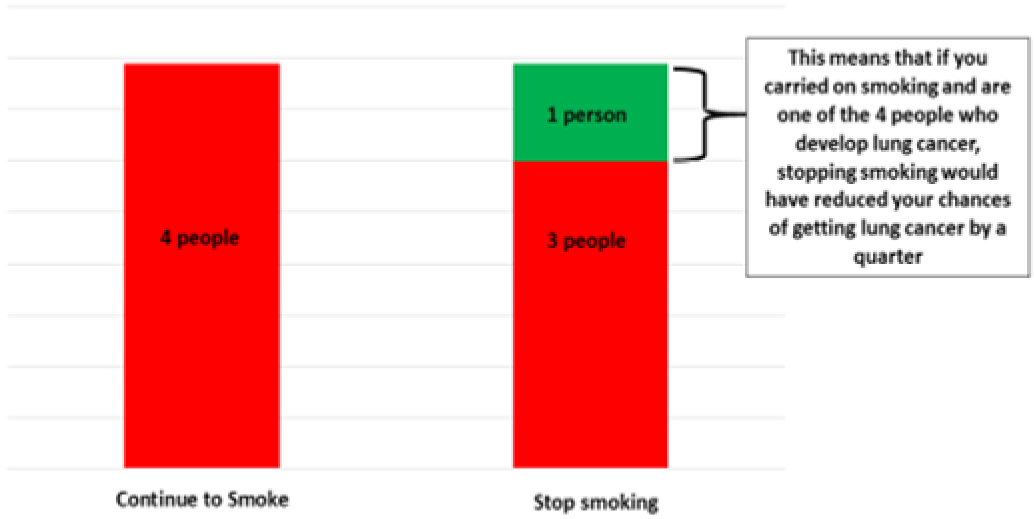

### Example 10

**'Based on the information we have collected about you, these pictures show the number of people like you out of 50 who will develop lung cancer if they continue to smoke…'**

**…In 2 years time**

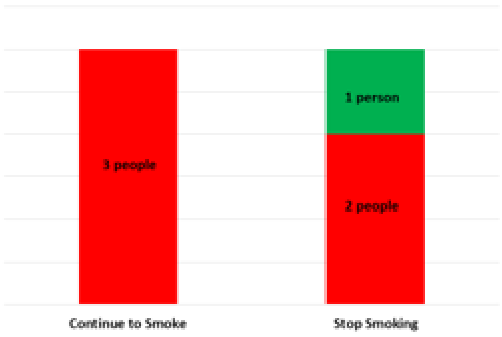


**…In 5 years time**

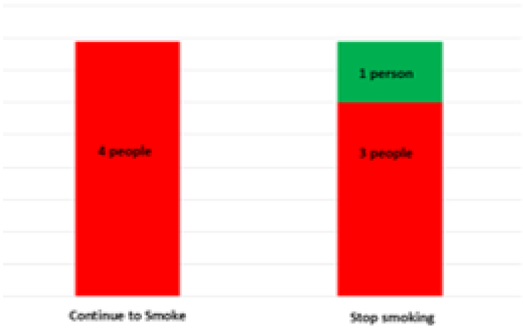


**…In 10 years time**

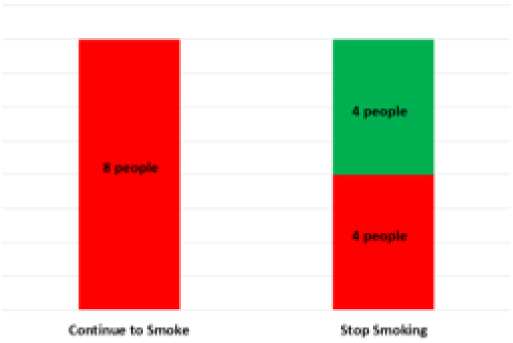

#### Examples provided in Focus Group 2

**Example 1**

**‘Based on information we have collected about you, we have worked out how stopping smoking would affect your specific risk of developing lung cancer in future. If 50 people like you continue to smoke, 4 people will develop lung cancer in 5 years’ time. If 50 people like you stopped smoking today, 3 people will develop lung cancer in 5 years’ time. That means out of 50 people like you, 1 fewer person will develop lung cancer in 5 years’ time if they stop smoking’**

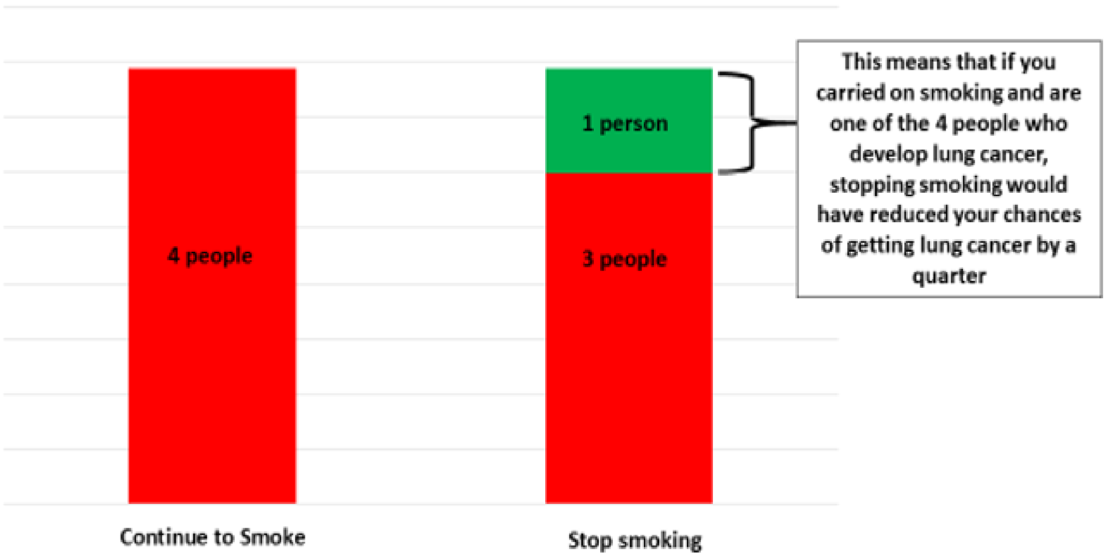

### Example 2

**'Based on the information we have collected about you, these pictures show the number of people like you out of 50 who will develop lung cancer if they continue to smoke…'**

**…In 2 years time**

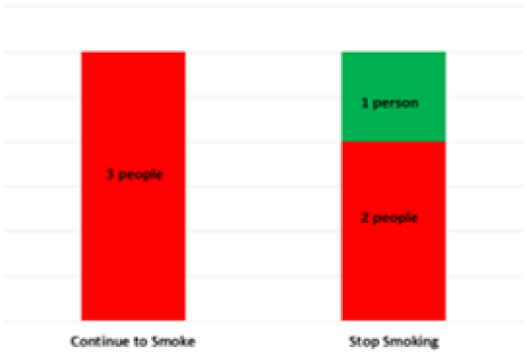


**…In 5 years time**

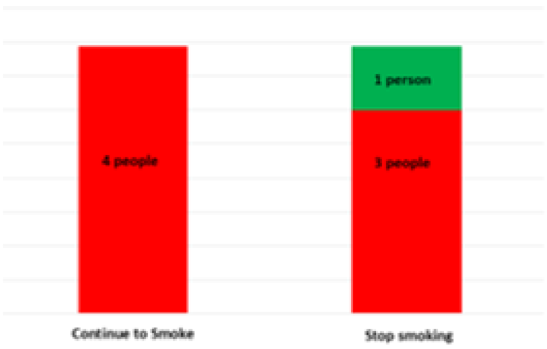


**…In 10 years time**

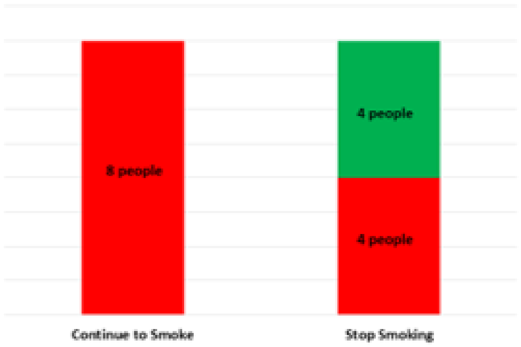


**…in their lifetime**

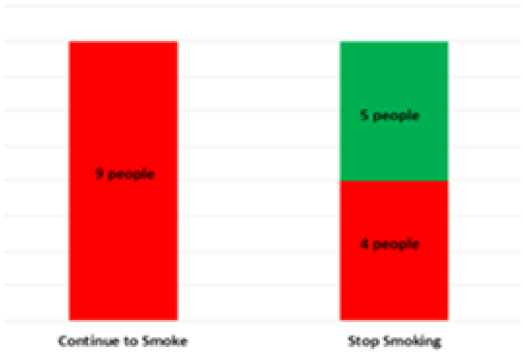

#### <u>Example 3</u>

**'Both of these pictures are your lungs. The darker parts on the picture on the left show the damage in your lungs because of smoking. The picture on the right shows healthy parts of your lung. These healthy parts have not yet been damaged by smoking.'**

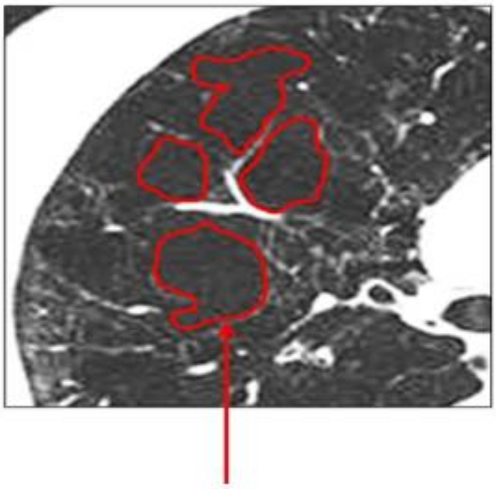

**Areas of damaged lung from smoking**

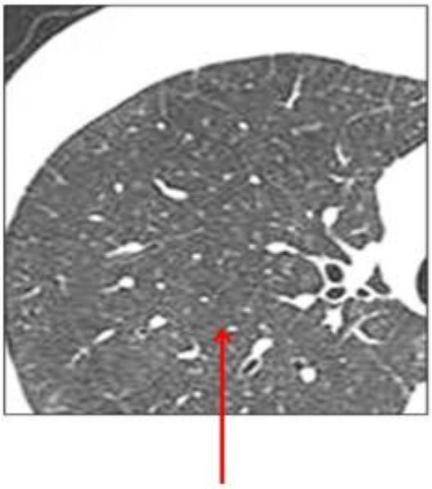

**Healthy parts of your lung that have not been damaged from smoking**

#### Example 4

**'The picture on the left is your lung. The darker parts on your lung have been damaged because of smoking. The picture on the right is a drawing of a lung. On this drawing, some parts have been damaged by smoking, and other parts are healthy.'**

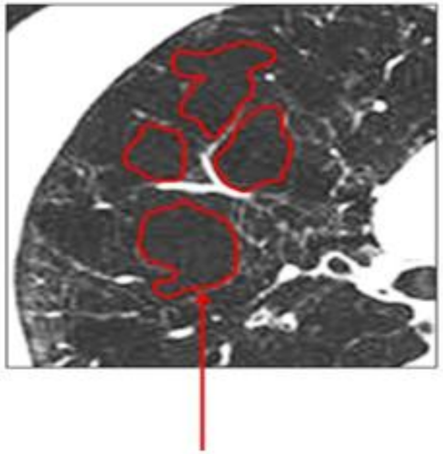

**Areas of damaged lung from smoking**

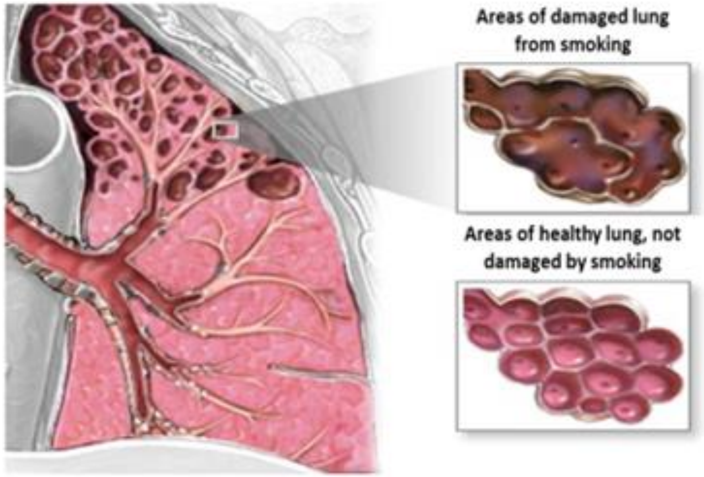

**Example 4a**

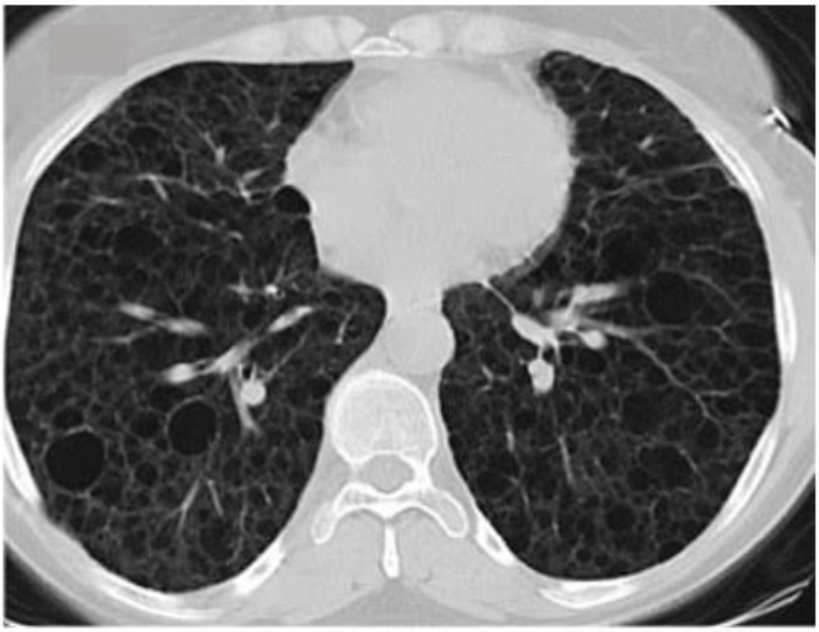

**Example 4b**

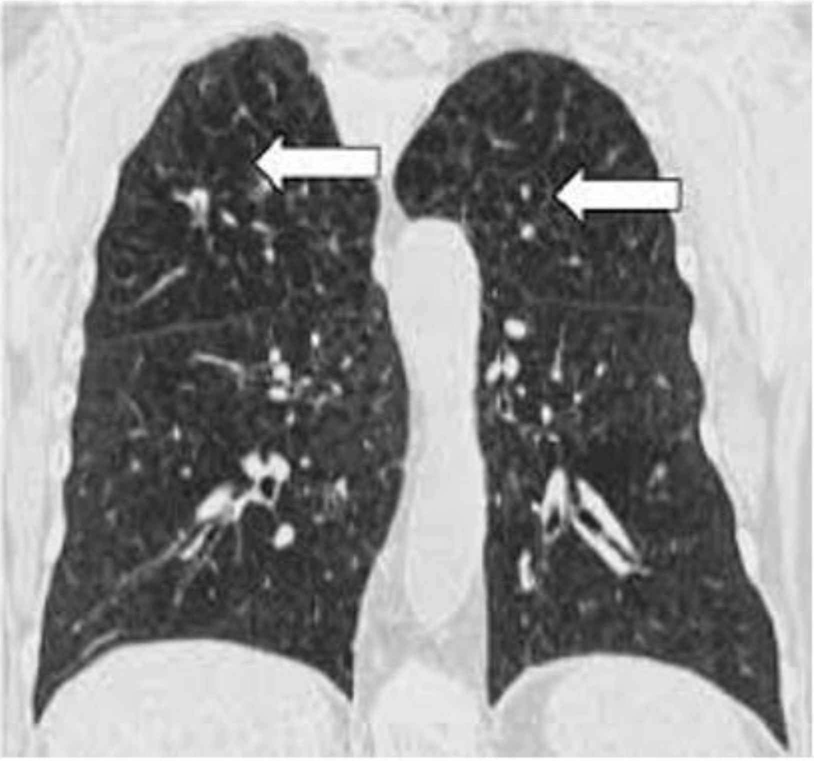

**Example 5**

**‘We can work out how old someone’s lungs are based on the information about them. For example, even though someone is actually 55 years old, because of various things like how much they smoke, their lung age might be 72 years old’**

#### Example 6

'The picture on the left is your heart. The picture on the right is a heart from a non-smoker. The yellow or white parts on your heart mean that the arteries going to your heart are more narrow than those of non-smokers heart. This is because smoking can cause narrowing of the artery going to the heart.'

Your heart

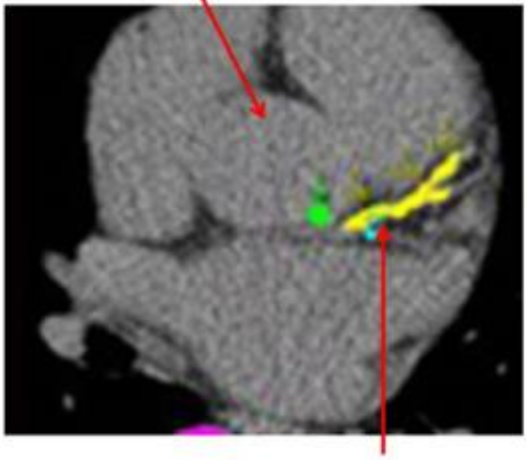

Yellow shows narrow arteries to the heart because of smoking

A normal heart

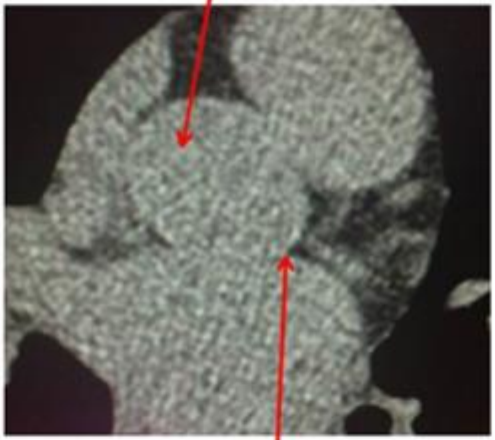

No narrowing of the arteries to the heart

Representative picture of your heart

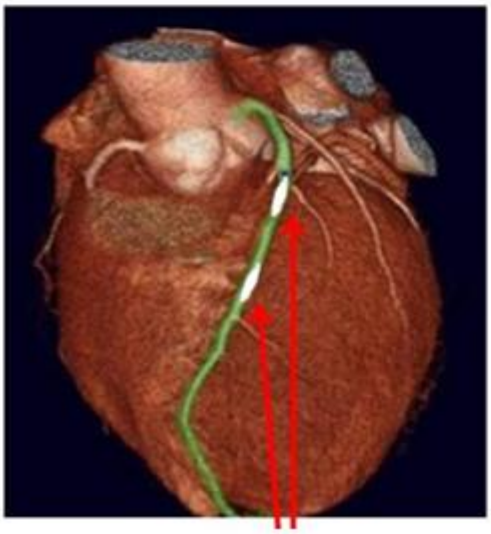

White shows narrow arteries to the heart because of smoking

A normal heart

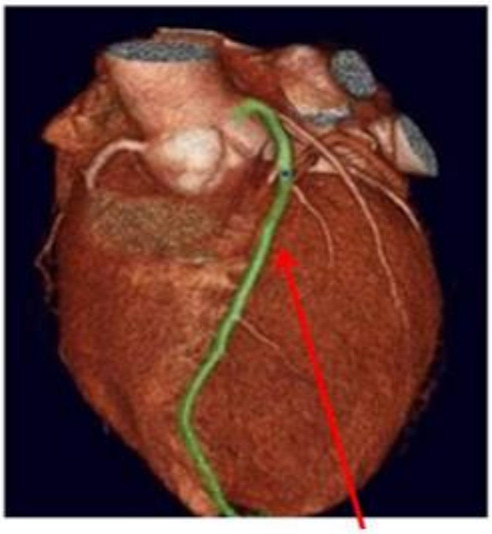

No narrowing of the arteries to the heart

### <u>Example 7</u>

**'The picture on the left is your heart. The white parts on your heart mean that the arteries going to your heart are narrower than those of non-smokers heart. This is because smoking can cause narrowing of the artery going to the heart. The picture on the right shows the difference between a normal and narrow artery.'**

White areas show narrow arteries because of smoking

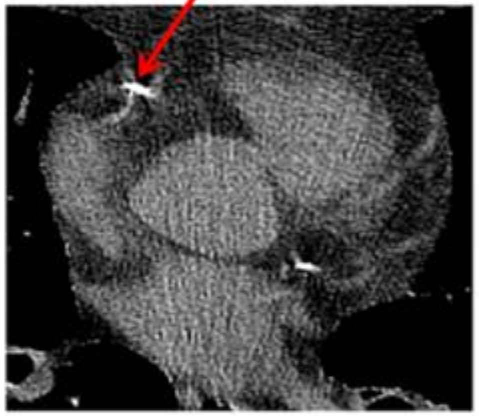

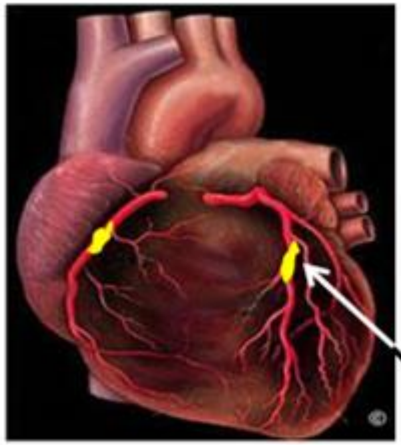

Yellow areas in your heart are narrow because of smoking

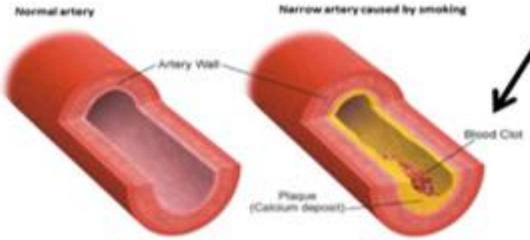

#### Focus Group 3

**Example 1**

**'Based on information we have collected about you, we have worked out how stopping smoking would affect your specific risk of developing lung cancer in future. If 50 people like you continue to smoke, 4 people will develop lung cancer in 5 years' time. If 50 people like you stopped smoking today, 3 people will develop lung cancer in 5 years' time. That means out of 50 people like you, 1 fewer person will develop lung cancer in 5 years' time if they stop smoking'**

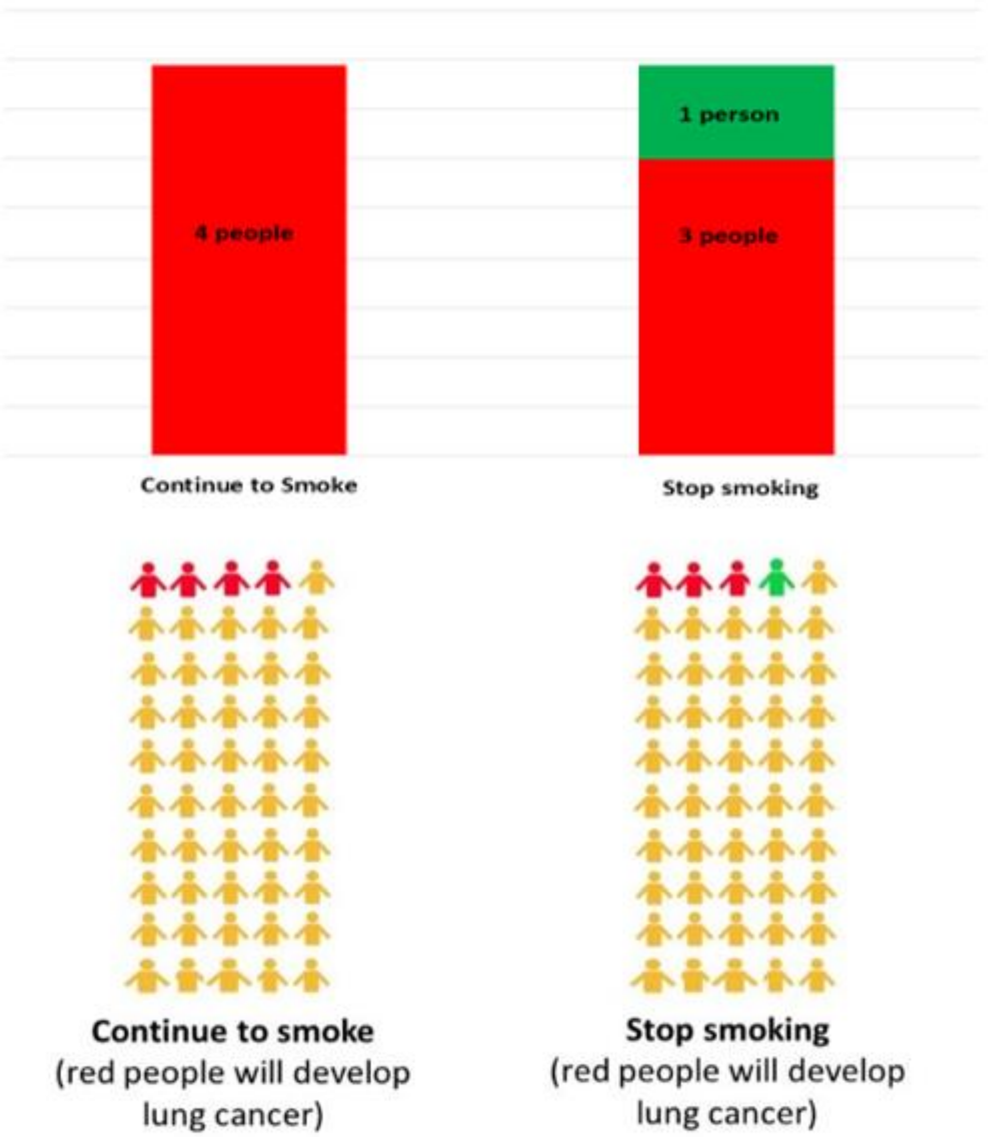

#### Example 2

**'Based on information we have collected about you, we have worked out how stopping smoking would affect your specific risk of developing lung cancer in future. If 50 people like you continue to smoke, 4 people will develop lung cancer in 5 years' time. If 50 people like you stopped smoking today, 3 people will develop lung cancer in 5 years' time. That means out of 50 people like you, 1 fewer person will develop lung cancer in 5 years' time if they stop smoking'**

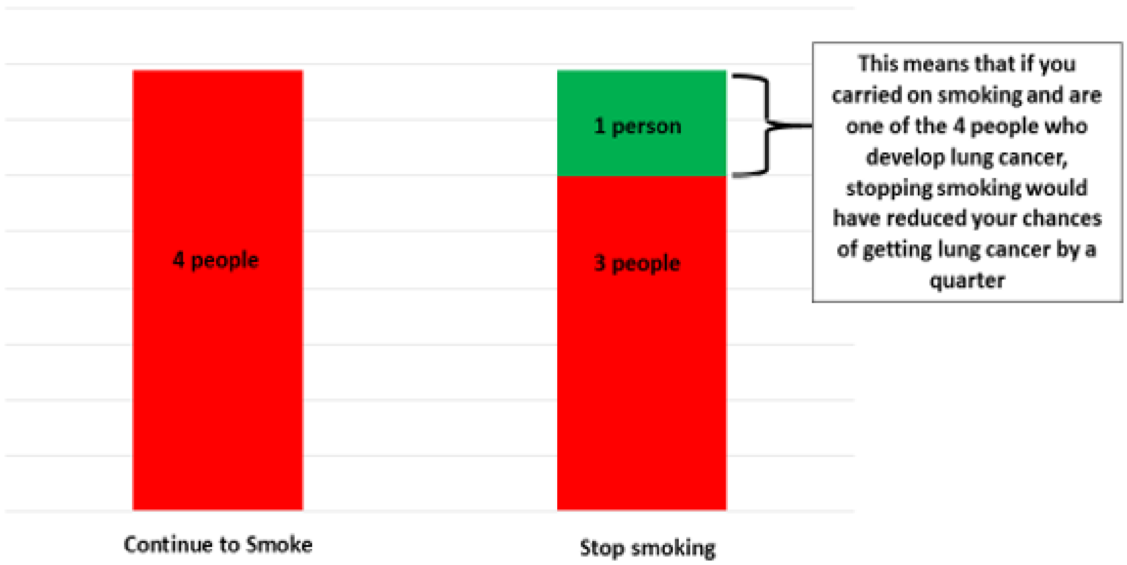

### Example 3

'Based on the information we have collected about you, these pictures show the number of people like you out of 50 who will develop lung cancer if they continue to smoke…'

...In 2 years time

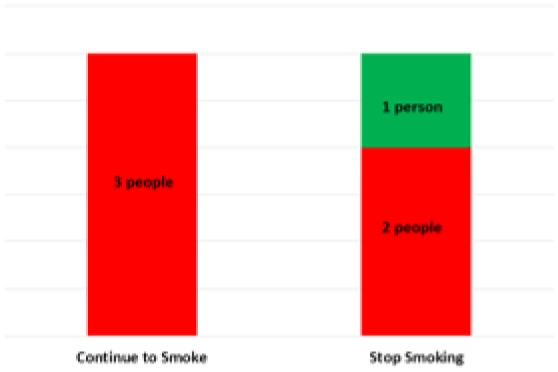


...In 5 years time

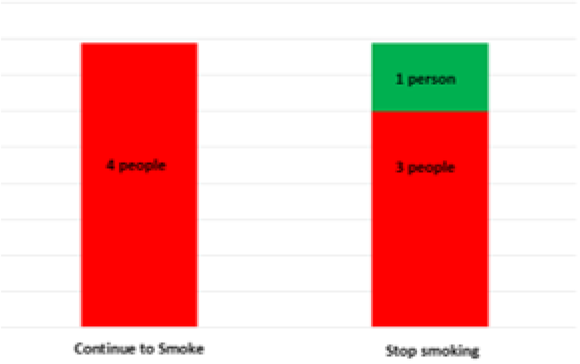


...In 10 years time

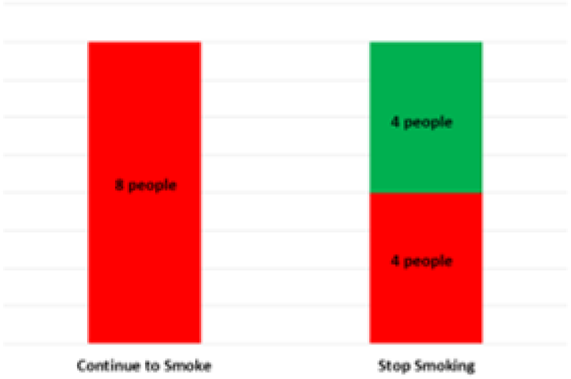

#### Example 4a

'The picture on the left is your lung. The picture on the right is a lung from a non-smoker. You can see that some parts of your lung are darker compared to the non-smokers lung. These darker parts are where the lung has been damaged because of smoking.'

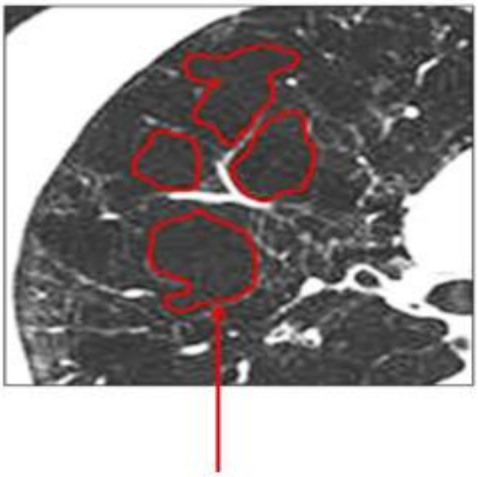

Areas of damaged lung from smoking

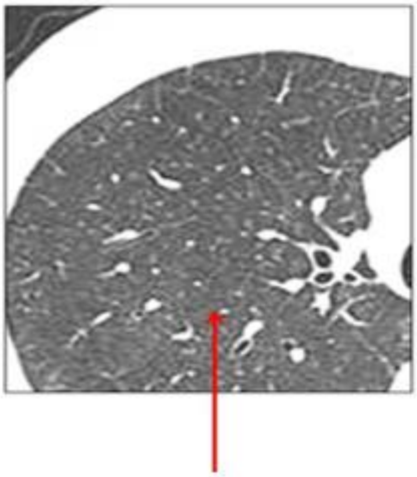

A healthy lung that has not been damaged by smoking

#### Example 4b

'Both of these pictures are your lungs. The darker parts on the picture on the left show the damage in your lungs because of smoking. The picture on the right shows healthy parts of your lung. These healthy parts have not yet been damaged by smoking.'

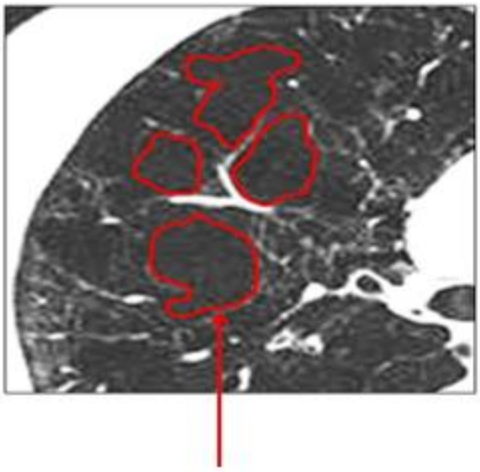

Areas of damaged lung from smoking

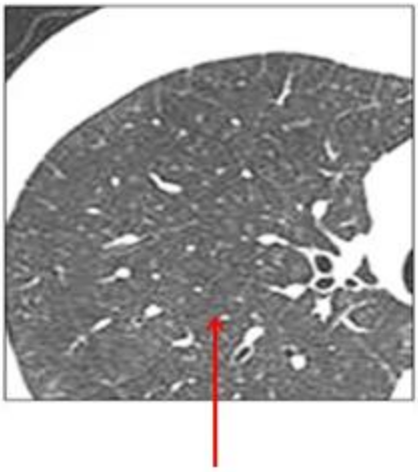

Healthy parts of your lung that have not been damaged from smoking

**Example 5**

**‘We can work out how old someone’s lungs are based on the information about them. For example, even though someone is actually 55 years old, because of various things like how much they smoke, their lung age might be 72 years old’**

#### Example 6

'The picture on the left is your heart. The picture on the right is a heart from a non-smoker. The yellow or white parts on your heart mean that the arteries going to your heart are more narrow than those of non-smokers heart. This is because smoking can cause narrowing of the artery going to the heart.'

Your heart

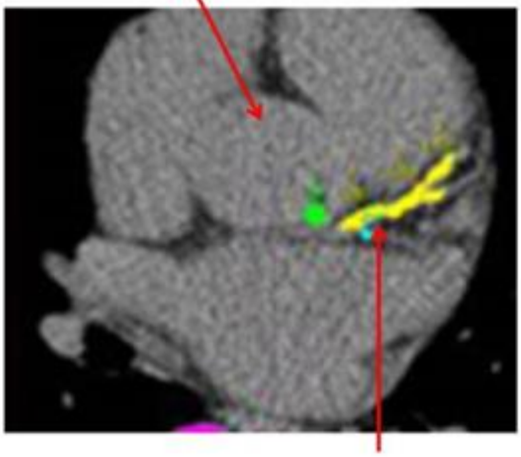

Yellow shows narrow arteries to the heart because of smoking

A normal heart

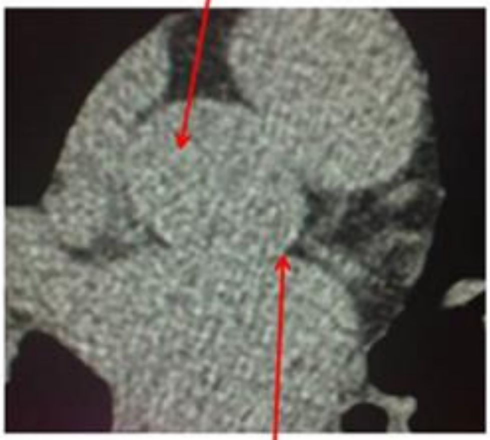

No narrowing of the arteries to the heart

**Representative picture of your heart**

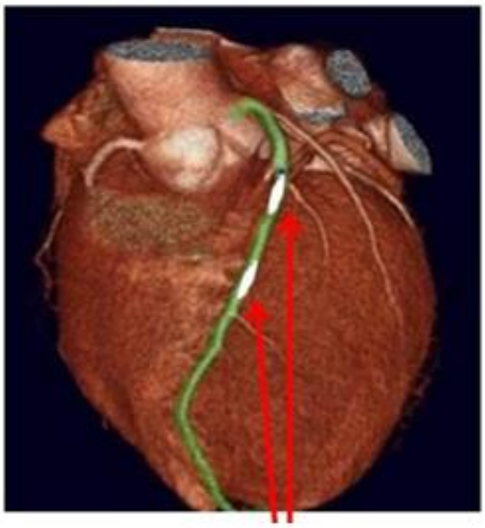

Yellow shows narrow arteries to the heart because of smoking

**A normal heart**

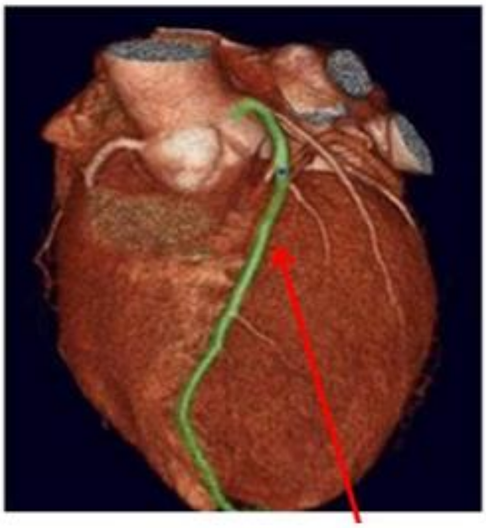

No narrowing of the arteries to the heart
