## Additional File 4 for "Co-development of an evidence-based personalised smoking cessation intervention for use in a lung cancer screening context"

Yorkshire Stop Smoking Study

Yorkshire Cancer Research

### Your scan results

University of Nottingham UK | CHINA | MALAYSIA

UNIVERSITY OF LEEDS

CARDIFF UNIVERSITY PRIFYSGOL CAERDYDD

UCL

NHS The Leeds Teaching Hospitals NHS Trust

### Your heart

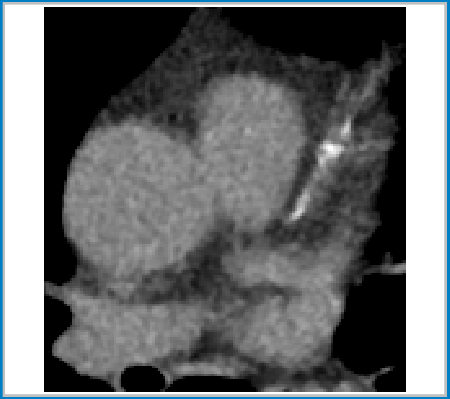

This is a picture of **your heart** taken from your CT scan.

The white areas show parts of the arteries going to your heart that have become hard or narrow.

Smoking is associated with the hardening and narrowing of the heart arteries. This can lead to problems including heart attacks.

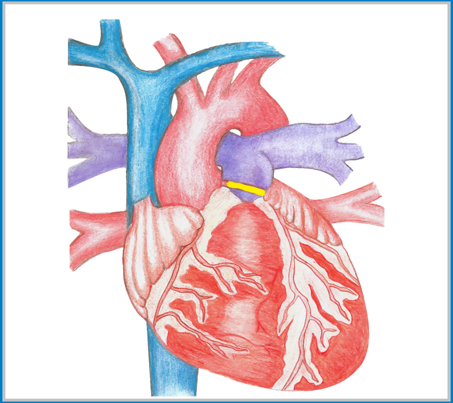

This drawing shows what your heart looks like inside.

The yellow parts show roughly where the arteries going to your heart have become hard or narrow.

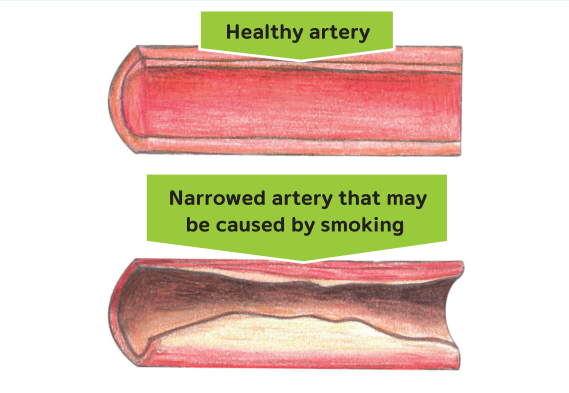


These are drawings of what healthy and narrowed arteries look like.

By stopping smoking, you can reduce the chance of the arteries to your heart becoming hardened or narrowed and lower your risks of problems like heart attacks.

### Your lungs

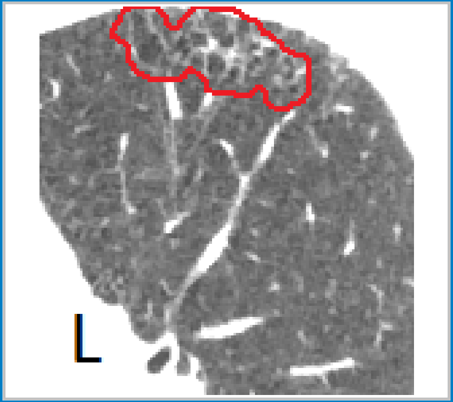


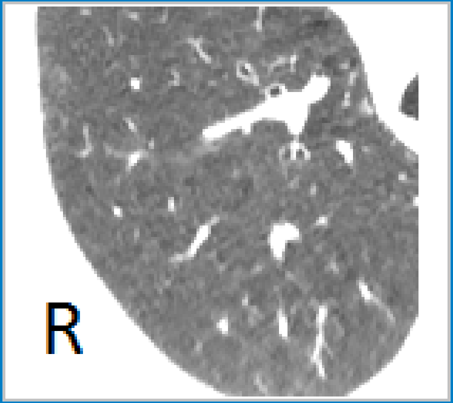


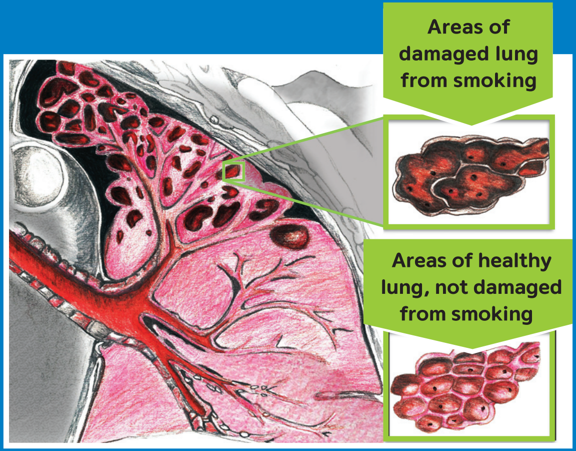


This is a picture of **your lung** taken from your CT scan.

The parts circled in red are areas of your lung that have been damaged by smoking.
This is called **'emphysema'**.

This is another picture of **your lung** taken from your CT scan.

This is a healthy part of your lung that has not been damaged by smoking.

You can keep these parts of your lung healthy if you stop smoking today.

This is a drawing of what your lungs look like inside.

The darker parts are the areas damaged from smoking (like the first picture).

The lighter parts are healthy areas not damaged by smoking (like the middle picture).

Stopping smoking will stop the healthy parts of your lungs from getting damaged.

### How stopping smoking will help your health

**After 20 minutes**
- Your heart rate goes back to normal.

**After 8 hours**
- Nicotine and carbon monoxide (a poisonous gas produced when smoking) in your blood goes down by half.
- Your oxygen levels go back to normal.

**After 2 days**
- You will not have any carbon monoxide left in your body.
- Your lungs will be clearer.
- You will be able to taste and smell better.

**After 3 days**
- You will fi nd it easier to breathe.
- You will have more energy and walking will be easier.

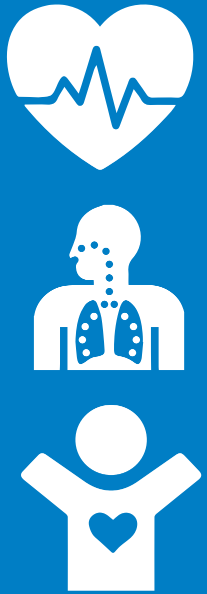

**After 2 to 12 weeks**
- Your blood will fl ow better around your body.
- This means more oxygen can get to important parts of your body.

**After 3 to 9 months**
- Your coughing and breathing will get better.
- Your lungs will start to work better - they can improve by up to 10%.

**After 1 year**
- Your risk of getting heart disease will go down to about half that of a person who is still smoking.

**After 10 years**
- Your risk of getting lung cancer will go down to about half that of a person who is still smoking.

**After 15 years**
- Your risk of having a heart attack will go down to the same as someone who has never smoked.

#### For more information get in touch with:

**Yorkshire Stop Smoking Study Project Manager**
Rebecca Thorley 0115 823 1361

**One You Leeds**
0800 169 4219

**Leeds Lung Health Check Clinical Team**
0113 392 6688

#### For any queries related to your smoking call:
