## Additional File 5 for "Co-development of an evidence-based personalised smoking cessation intervention for use in a lung cancer screening context"

**YESS SCP scripts to accompany intervention booklet**

| | AIMS | APPROACH/SCRIPTS |
|---|---|---|
| **1. REMIT OF USUAL CARE FOR SMOKING CESSATON ADVISOR (BOTH INTERVENTION AND CONTROL GROUPS)** | | |
| | Behavioural support | N/A |
| | NRT/medication | N/A |
| | MI/self-efficacy for quitting | N/A |
| **2. INTERVENTION BOOKLET** | | |
| ***Brief for SCPs*** | Increase self-efficacy and response efficacy through personalisation of results/risk<br><br>Ensure salience and credibility of results/intervention<br><br>Manage potential for both anxiety and over-reassurance following results<br><br>Ensure the risk information and contents of the booklet are understood<br><br>Set out expectations from the start for results feedback (i.e. that the smoking cessation practitioner will not be able to answer clinical questions about results) | Approach:<br>- Encourage, motivate and empathise<br>- Emphasise immediate/short-term benefits of quitting regardless of scan result and importantly, for any age<br>- Build confidence in ability to quit (covered by usual care for all)<br>- Be sympathetic about, and acknowledge, the difficulties of quitting<br>- Relate results/risk and proposed benefits directly to the individual<br>- Present YESS and YLST as professional, expert services<br>- Check understanding/comprehension<br>- Explain role of SCP: that they are not a medical doctor and therefore the limits to the clinical advice that can/will be given by YESS smoking cessation practitioners. Highlight that there is a phone number on the booklet if they have any queries that they wish to discuss with the YLST clinical team.<br>- Manage impact of results on risk perceptions – appropriate reassurance |
| ***Introduction to booklet*** | Emphasise salience from outset (these are *your* images)<br><br>Encouragement<br><br>Expectation setting | This booklet gives information about your CT scan results. This includes pictures from your own CT scan that may show damage caused by smoking. This is to show how your smoking has affected your lungs and heart, but importantly, how much of a positive change you can make to your future health. Whatever your age and whatever your CT scan shows, stopping smoking will reduce your risk and improve your health. Just by being here today, you've already taken a really important step and you now have the right support to give you every chance of success. |

| | | |
|---|---|---|
| ***'Your lungs' page*** | Describe lung images (bold text taken directly from booklet)<br><br>Clarify continued risks of smoking in context of CT result by using lay explanation of how smoking causes emphysema and how emphysema affects the participant's health<br><br>Increase response efficacy by explaining how stopping smoking has immediate benefits of limiting further damage (for those with damage) or preventing damage (for those without damage).<br>Manage/reduce potential for over-reassurance among those with 'no' lung damage | <u>**Script for participants with 'some' lung damage**</u><br>The first picture is a **picture of your own lung taken from the CT scan. The parts circled in red are areas of your lung which have been damaged by smoking. This is called emphysema.** Emphysema means there is damage to the tiny air sacs in your lungs, which has been caused by tobacco smoke. This damage causes these air sacs to join together and trap pockets of air inside the lungs. Over time, small areas of the lung are destroyed, so that instead of working lung, there are empty holes. As more of the lung gets damaged, this causes difficulties with breathing.<br><br>This is another picture from your CT scan, which shows a **healthy part of your own lung that has not been damaged by smoking.** If you continue to smoke, then this part of your lung may become damaged in the same way. But if you stop smoking today then this part of your lung will stay healthy. It really is never too late to make positive changes. You will immediately be protecting the healthy air sacs in your lungs and keeping as many of them working as possible. After just three days, you should start to feel better in yourself.<br><br>**This is a drawing of what a lung looks like inside. The darker parts are the areas damaged from smoking like in the first picture** of your lung. **The lighter parts are healthy areas not damaged by smoking like in the middle picture** of your lung. |
| | | <u>**Script for participants with 'extensive' lung damage: Scenario A**</u><br>[If still able to show a picture of 'healthy' part of own lung, then same as above] |
| | | <u>**Script for participants with 'extensive' lung damage: Scenario B**</u><br>[If the 'healthy' part of lung is a 'less damaged' part of the lung].<br><br>The first picture is a **picture of your own lung taken from the CT scan. The parts circled in red are areas of your lung which have been damaged by smoking. This is called emphysema.** Emphysema means there is damage to the tiny air sacs in your lungs which has been caused by tobacco smoke. This damage causes these air sacs to join together and trap pockets of air inside the lungs. Over time, small areas of the lung are destroyed, so that instead of working lung, there are empty holes. As more of the lung gets damaged, this causes difficulties with breathing. |

| | | |
|---|---|---|
| | | This is another picture from your CT scan which shows a **part of your own lung that has less damage from smoking.** If you stop smoking today, then you will immediately be protecting these air sacs in your lungs and keeping them working as well as possible. This will help stop or slow down any further damage so that your breathing does not get worse. It really is never too late to make positive changes. After just three days, you should start to feel better in yourself.<br><br>This is a drawing of what a lung looks like inside. The darker parts are the areas with more damage from smoking like in the first picture of your lung. The lighter parts are areas with less damage from smoking like in the middle picture of your lung. |
| | | <u>**Script for participants with 'no' lung damage**</u><br>The first picture is a **picture of your own lung taken from your CT scan.**<br><br>The second picture is of a smoker's lung who is older than you. **The parts circled in red are areas of the lung which have been damaged by smoking. This is called emphysema.** Emphysema means there is damage to the tiny air sacs in the lungs which has been caused by tobacco smoke. This damage causes these air sacs to join together and trap pockets of air inside the lungs. Over time, small areas of the lung are destroyed, so that instead of working lung, there are empty holes. As more of the lung gets damaged, this causes worsening difficulties with breathing. While there is no visible damage to your lungs yet, if you continue to smoke, then this part of your lung is likely to become damaged in the same way. But if you stop smoking today then this part of your lung will stay healthy. It really is never too late. You will immediately be protecting the healthy air sacs in your lungs and keeping as many of them working as possible. After just three days, you should start to feel better in yourself.<br><br>This is a drawing of what a lung looks like inside. The darker parts are the areas with more damage from smoking like in the second picture of an older smokers' lung. The lighter parts are areas with less damage from smoking like in the first picture of your lung. |

| | | |
|---|---|---|
| ***'Your heart' page*** | Describe heart images (bold blue text taken directly from booklet)<br><br>Clarify continued risks of smoking in context of CT result by explaining that smoking is one likely cause of their artery narrowing and hardening. Set this in the context of other causes so as not to over-exaggerate role of smoking, which participants may discredit.<br><br>Manage/reduce potential for over-reassurance among those with 'no' heart damage | **Script for participants with 'some' heart damage**<br>The first picture is a **picture of your own heart taken from your CT scan. The white parts of your scan picture are parts of the arteries going to your heart that have become narrow or hard, which is called "hardening of the arteries".** It is harder for blood to flow through these arteries, which means that your heart can't always get the supply of oxygen it needs to work properly - increasing your chance of problems like angina or heart attacks in the future. There are lots of factors which may have caused this hardening and narrowing, but your smoking is likely to be one of the most important of them.<br><br>This second picture **shows what your own heart looks like inside. The white parts show roughly where the arteries going to your heart have become hard or narrow.** You can see that there are also healthy parts of your arteries too, which haven't been damaged by your smoking yet.<br><br>**These are drawings of what healthy and narrowed arteries look like. By stopping smoking, you can** keep your arteries as healthy as possible, which is really important at your age. This will reduce any further damage from your smoking. Your heart will start to benefit from quitting very quickly. In just two weeks from now, your blood flow will start to improve. This means a healthier heart, which is able to get a better supply of oxygen to help it work properly. It also means you'll have a lower chance **of problems like heart attacks** or angina**.** In just one year from now, your risk of heart disease will go down to about half that of a smoker. |
| | | **Script for participants with 'extensive' heart damage**<br>[If still able to show picture of 'healthy' part of own heart, then same as above] |
| | | **Script for participants with 'no' heart damage**<br>The first picture is a **picture of your own heart taken from your CT scan.**<br><br>This second picture shows a heart that has been damaged by smoking. **The white parts are where the arteries going to the heart have become hard or narrow.** It is harder for blood to flow these arteries, which means that the heart can't always get the supply of oxygen it needs to work properly - increasing the chance of problems like angina or heart attacks in the future. There are lots of factors which |

| | | |
|---|---|---|
| | | may have caused this hardening and narrowing, but smoking is one of the most important of them.<br><br>**These are drawings of what healthy and narrowed arteries look like. By stopping smoking, you can** keep your arteries as healthy as possible and prevent any damage. This is really important at your age. While there is no visible damage to your arteries yet, as a smoker you are at high risk of this damage in the future. Your heart will start to benefit from quitting very quickly. In just two weeks from now, your blood flow will start to improve.<br><br>This means a healthier heart, which is not having to work as hard. It also means you'll have a lower risk **of problems like heart attacks.** In just one year from now, your risk of heart disease will go down to about half that of a smoker. |
| ***How stopping smoking with help your…*** | **HEALTH** (bold blue text taken from booklet)<br><br>Increase response efficacy by emphasising the short-term benefits of stopping smoking and relating this directly to the individual; helping them envision how their health will improve over the first year (focussing less on the longer term). Use lay explanations to show how these health benefits accumulate and work to achieve significant reductions in risk over the following years.<br><br>Emphasise relevance for older age from the start (and continually if needed). | This page shows just how much, and how quickly, stopping smoking will improve your health, and at any age no matter how long you have smoked. The benefits start after just 20 minutes.<br><br>Your health will start to improve just 20 minutes after you stop smoking. That's when your heart rate will **go back to normal**.<br><br>After eight hours, the amount of **nicotine and carbon monoxide in your blood will go down by half**. Nicotine is the chemical in tobacco that makes smoking addictive. Carbon monoxide is a poisonous gas in cigarette smoke that replaces the oxygen in your blood stream. Breathing in carbon monoxide can lead to heart disease. But after just eight hours of not smoking, your levels of both these chemicals will reduce by half. After two days, your body will have got rid of all the carbon monoxide from smoking and your lungs will be clearer. You should also notice that your taste and sense of smell are better. After three days, you may start to find it **easier to breathe, you may have more energy**, and you could find moving around feels a little easier. |

| | | |
|---|---|---|
| | | Over the next two to 12 weeks, the health benefits will continue. By this point, your blood will be **flowing better around your body**. More and more oxygen will be **getting to the important parts of your body**.<br><br>Over the following three to nine months, you should really start to notice a difference with your lungs. Your breathing and coughing will have got better. This is because your lungs will be able to work much better. Your lung function should have improved **by up to 10%.**<br><br>Over the following 10 to 15 years, **your risk of having a heart attack will go down to the same as someone who has never smoked**. Your risk of lung cancer will be about half that of a person who is still smoking. As it will begin to fall steadily from the day you quit due to all the positive changes that will start happening in your body straight away.<br><br>**<u>Extra text for participants with no evidence of heart disease</u>**<br>All of these positive changes in your body mean that a year from today, **your risk of getting heart disease will go down to about half that of a person who is still smoking.** |
| | **WEALTH** (bold blue text taken from booklet)<br><br>Describe personalised financial savings from quitting | You know better than me that smoking is expensive. From the information you gave at your Lung Health Check, we've worked out how much better off you will be financially. **Based on the [number] cigarettes you are currently smoking per day, stopping smoking would save you [number] pounds per week, [number] pounds per month, and [number] pounds per year.** This means a saving of [**number] pounds in the next five years**. |
| **3. SPECIFIC RESPONSES** | | |
| | Managing anxiety about the extent of smoking damage, including questions about this not being reversible. | Approach:<br>- Listen to and empathise with concerns and distress<br>- Acknowledge concerns about damage (avoid temptation to play down damage to reduce distress)<br>- Reassure that stopping smoking will have a positive impact by preventing further damage / maintaining healthy areas. Explain that damage is not |

| | | |
|---|---|---|
| | | reversible but always end positively by reassuring that the benefits of stopping smoking begin immediately. Build the participant's sense of control over their health by praising them for the positive step they are taking in quitting.<br>- Re-iterate that YESS will offer them the best possible support for quitting and build self-efficacy/confidence<br>- Reassure them that they are receiving expert respiratory care as part of YLST and are proactively looking after their health<br>- Signpost for support from YLST clinician if necessary (limited capacity and not routine for all) and perhaps book them with an appointment for a telephone consultation before the session ends) |
| | Managing anxiety and concerns about disclosure of emphysema and coronary artery calcification | Approach<br>- Listen to and empathise with concerns and distress<br>- Explain that YESS will offer them the best possibly support with quitting but clinical questions are best answered by YLST clinicians<br>- Signpost for support from YLST clinician (perhaps book them with an appointment before the session ends and provide with helpline details) |
| | Manage scepticism about the role of smoking in existing (for those with damage) or future (for those without damage) heart disease | Approach<br>- Acknowledge, and be able to explain, the role of other risk factors for heart disease (i.e., diet) while still emphasising the role that smoking plays, and importantly the role that stopping smoking could play in preventing disease progression |
| **4. CLOSING SESSION** | | |
| | Positive close | Reiterate key messages set out at the beginning e.g. emphasise immediate benefits of quitting, best thing they can do for their health regardless of scan result/age ("it's never too late"), and expert support is on hand all the way, etc. |
| | Further safety net against potential over reassurance for those with clear results | Reiterate to those with clear results that the way to keep their lungs/heart healthy is to stop smoking and while it's good news, their risk of future damage is high |
