## Additional File 6 for "Co-development of an evidence-based personalised smoking cessation intervention for use in a lung cancer screening context"

Department of Behavioural Science and Health

UCL

### Yorkshire Stop Smoking Study (YESS)

### Intervention Training for SCPs

YORKSHIRE STOP SMOKING STUDY | LEEDS LUNG HEALTH CHECK |

Monday 3rd December

### Outline

- Intervention overview
  - Evidence-base for development
  - Special considerations for participant group

- Intervention components
  - Key elements of behavioural support
  - Intervention group vs. control group

- Intervention scripts
  - Guidance and standardisation
  - Variation in scenarios (i.e. results, quit stages)

### INTERVENTION OVERVIEW

### Background to intervention

- Evidence base for intervention development drew on:
  - Existing studies of personalised risk information
  - Online survey and focus groups to gauge preferences for format of visual presentation
  - PPI groups to review intervention (i.e. acceptability, understanding)

### Special considerations for screening-eligible

- Compared with general population:
  - Lower socioeconomic status (SES)
  - Lower health literacy and numeracy
  - Smoking-related morbidities
  - Clustering of cancer risk behaviours

- Compared with smoking population in general:
  - Higher nicotine dependence
  - Less success/willingness/confidence in quitting
  - Longer-term tobacco addiction (commonly since childhood)

### Intervention components

- Enhanced personalised risk information booklet
  - Images of own CT scan results
  - Healthy and unhealthy areas of lung and heart
  - Generic lung, heart and artery drawings
  - Immediate/short-term health benefits of stopping smoking

- Scripted advice for smoking cessation practitioners
  - Standardisation important for intervention fidelity
    - But doesn’t need to be verbatim
  - Enhance delivery/behavioural support of risk information

### INTERVENTION COMMUNICATION: KEY ELEMENTS OF BEHAVIOURAL SUPPORT

### Capability, opportunity and motivation

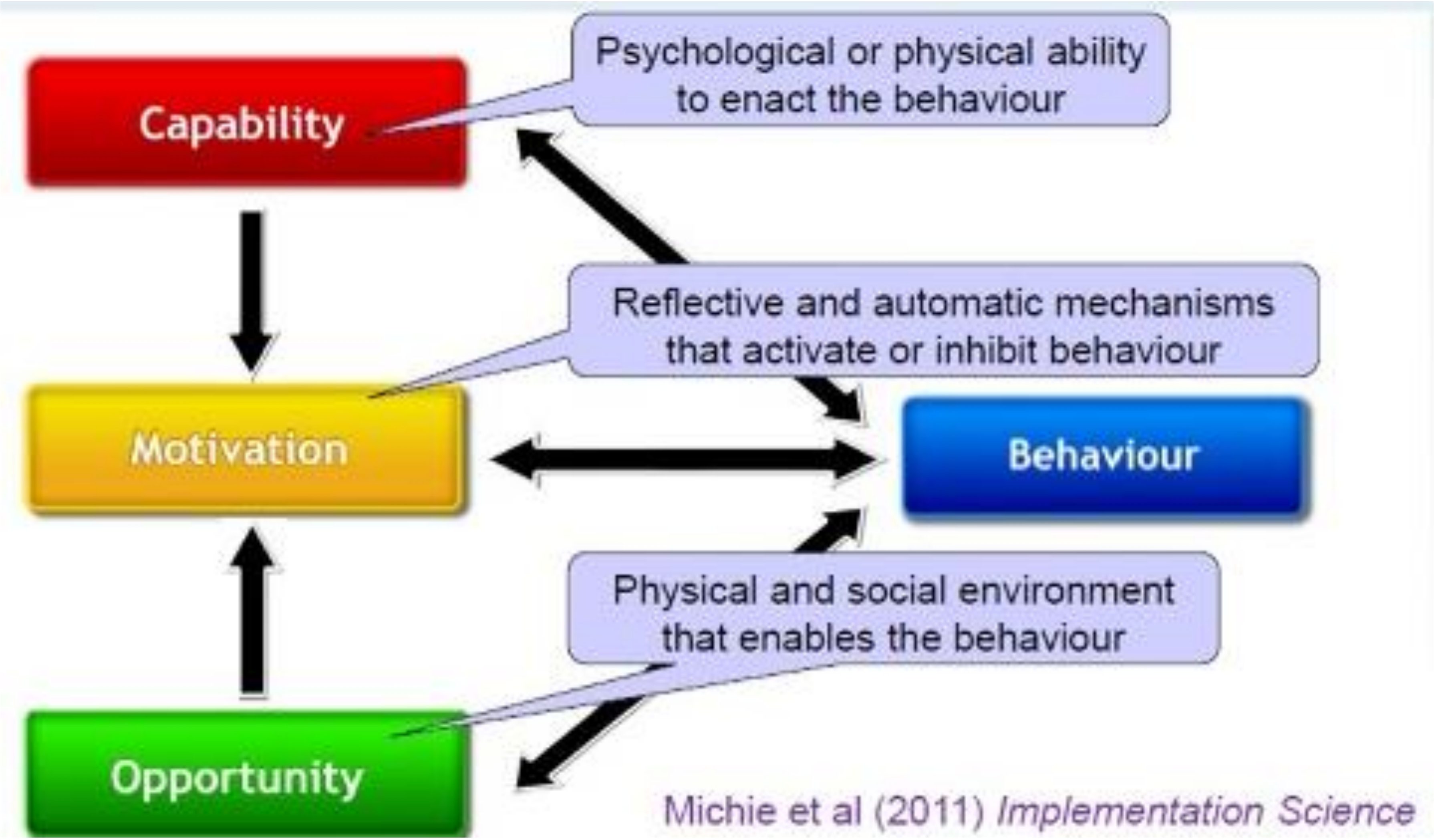


**The COM-B Model**

### Responses to risk information

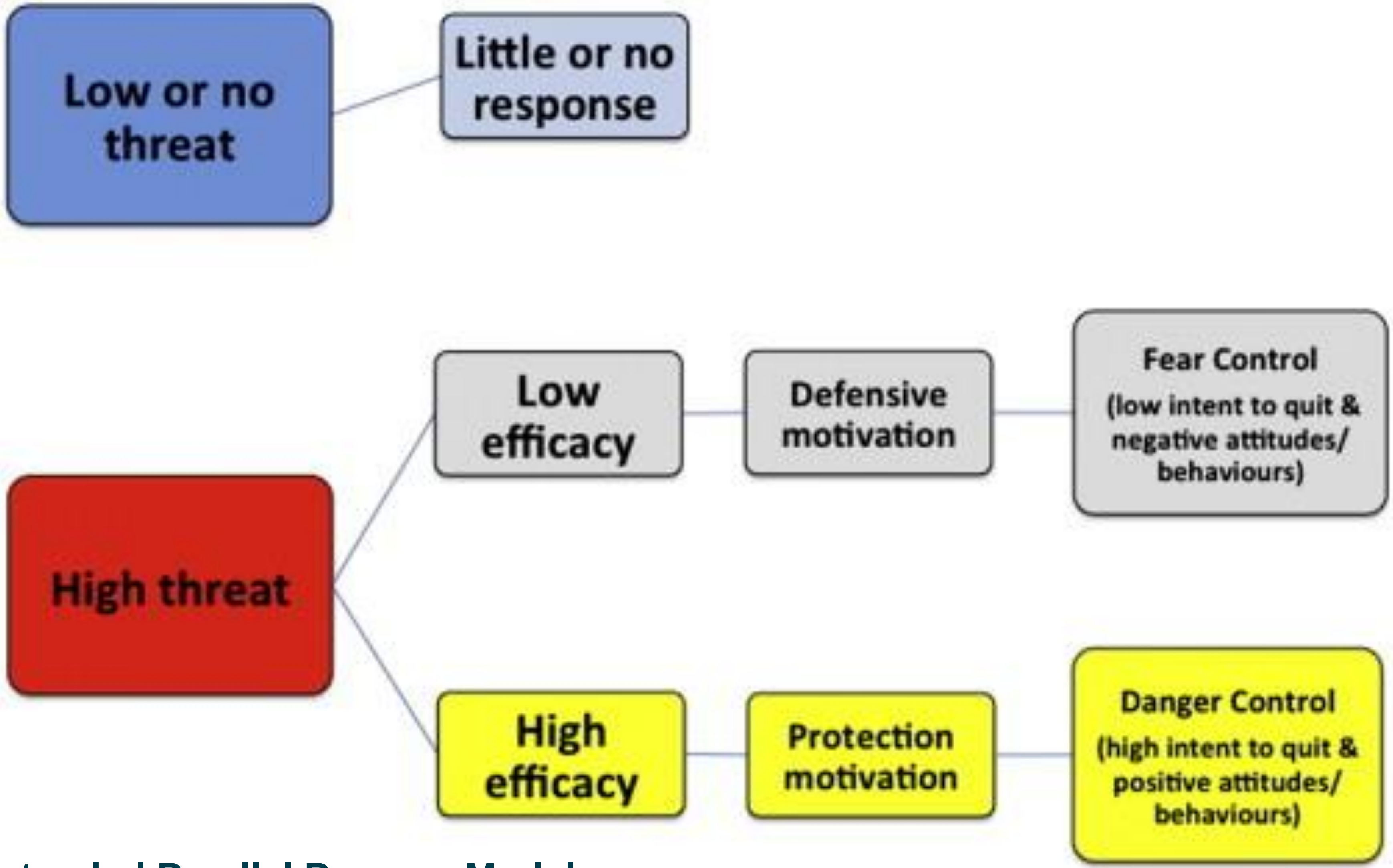


**Extended Parallel Process Model**

### Efficacy beliefs (extremely important!)

- Self-efficacy
  - An individual’s belief in their ability/capability to carry out a behaviour/goal successfully
  - Predicts whether people take up behaviours and persist with those which are effortful/difficult to achieve

- Response efficacy
  - An individual’s belief in whether the recommended behaviour will address the threat
  - i.e. will stopping smoking improve health/reduce health risks
  - They need to believe they will benefit

### Importance of emphasising benefits

*"I was quite prepared to be shocked [at the Lung Health Check], your lungs are in this condition. We've taken photographs. We've looked at it. You've got COPD and this is the level you're at with your breathing. To then say, look,* ***<u>if you hadn't have done that you'd be there and if you stop you can get there</u>****. I need to know the state I'm in no matter what that entails. Obviously, nobody wants to hear they've got cancer, but if I have bloody tell me. I want to know. Now, how are my lungs now, compared to if I'd never smoked? It's going to be pretty bad,* ***<u>but if I give up, how far back can I come</u>****. I know there's a certain amount of damage done that I'm never going to come back from, but it must improve if I stop.* ***<u>Where can I get back to, what can I aim at, an incentive today</u>*** *that this is how bad you are, you give up,* ***<u>you can maybe get to there</u>****…… as low as I am lung wise, that as low as my health is through smoking,* ***<u>show me some positives</u>****."*

**Interview study (in prep) with participants in Lung Screen Uptake Trial**

### Building efficacy beliefs

- Credibility of risk information and <u>you</u>

- Emphasise health benefits
  - At any age
  - Regardless of smoking duration

- Immediacy of health benefits
  - Bias toward short-term benefits (low SES and older age)

- Explain process of damage/benefit
  - Lay explanations of how smoking damages the lungs and heart
  - Processes are cumulative and ongoing (not black and white)

### Optimising how participants respond

- Objective: motivational impact of results for smoking cessation

- Manage anxiety about perceived risk
  - Acknowledge and listen to concerns
  - Present smoking cessation as proactive risk management
    - Building efficacy beliefs
  - Avoid temptation to downplay risk to lower someone’s distress

- Manage over reassurance about perceived risk
  - Clear results are good news but do not change future risk
  - Use as catalyst for change

### Style of communication

- Encouragement
- Empathy
- Check understanding (e.g., talk back)
- Reflective listening
- Non-verbal communication

### Different result scenarios

- Different possible results
  - Emphysema: none, some or severe
  - Coronary artery calcification: none or some
  - Comparison images vary depending on result
  - Scripted advice on slides is colour coded: green, amber, red
- Spectrum of different result combinations
  - From severe damage to mixed to no damage
  - Extremes have potential to cause anxiety or over-reassure

### Different quit stage/readiness scenarios

- Since Lung Health Check…
  - Quit and remained abstinent
  - Quit and relapsed
  - Ready to quit

- Adapt wording accordingly
  - i.e. if you continue to smoke vs. if you hadn't have stopped

- Recent relapse
  - Self-efficacy especially crucial

### INTERVENTION SCRIPTS: Introduction

### Preparatory conversation

- Credibility
  - Introduce YESS and YLST as professional, expert services
- Personalise
  - Emphasise from the outset these are <u>your</u> pictures
- Set expectations for results feedback
  - Will see own images and may see damage
  - Explain your role as a smoking cessation practitioner
  - Limits to clinical advice (highlight YLST number on booklet)
- Encourage

### INTRO SCRIPT

**PREPARE – PERSONALISE – ENCOURAGE**

"This booklet gives information about **your** CT scan results. This includes pictures from **your own** CT scan that may **show damage caused by smoking**. This is to **show how your smoking has affected your lungs and heart**, but importantly, **how much of a positive change you can make to your future health. Whatever your age and whatever your CT scan shows, stopping smoking will reduce your risk and improve your health**. Just by being here today, **you've already taken a really important step** and **you** now have the **right support** to give **you every chance of success**."

### INTERVENTION SCRIPTS:
### "Your lungs"

### Example for 'some' damage

#### Your lungs

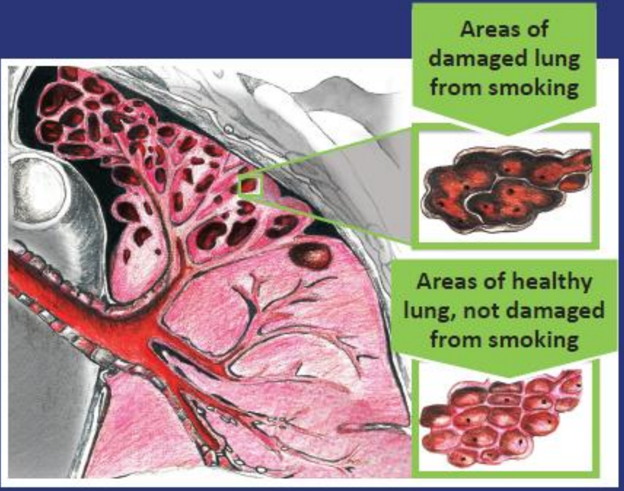


This is a picture of **your lung** taken from your CT scan.

The parts circled in red are areas of your lung that have been damaged by smoking.
This is called **'emphysema'**.

This is another picture of **your lung** taken from your CT scan.

This is a healthy part of your lung that has not been damaged by smoking.

You can keep these parts of your lung healthy if you stop smoking today.

This is a drawing of what your lungs look like inside.

The darker parts are the areas damaged from smoking (like the first picture).

The lighter parts are healthy areas not damaged by smoking (like the middle picture).

Stopping smoking will stop the healthy parts of your lungs from getting damaged.

### Presenting lung images

- Describe lung images
- Explain risk
- Clarify continued risk of smoking in context of result
  - Lay explanation of how smoking causes emphysema

- Increase response efficacy
  - Immediate benefits of preventing/limiting smoking damage

- Manage potential for over-reassurance

### YOUR LUNGS SCRIPT (picture 1)

***"The first picture is a picture of <u>your own</u> lung…***

***Severe or some damage: "…***taken from **<u>your</u>** CT scan. The parts circled in red are **<u>areas of your lung</u>** which have been **<u>damaged by smoking</u>**. This is called **<u>emphysema</u>**."

***Some damage:*** "…taken from **<u>your</u>** CT scan. The parts circled in red are **<u>areas of your lung</u>** which have been **<u>damaged by smoking</u>**. This is called **<u>emphysema</u>**."

***No damage: "…***taken from **<u>your</u>** CT scan."

### YOUR LUNGS SCRIPT (picture 2)

***"The second picture is…***

***Severe damage:*** *…* a part of your own lung that has **less damage** from smoking."

***Some damage:* …** *a* **healthy** part of your own lung that has not been damaged by smoking."

***No damage:* …** of a **smoker's lung who is OLDER than you**. The parts circled in red are areas of the lung which have been **damaged by smoking**. This is called **emphysema**."

### YOUR LUNGS SCRIPT (emphysema)

- Lay explanation of emphysema describing **<u>how</u>** smoking damages the lungs and that this **<u>process is cumulative</u>**

***"Emphysema means…***

*…there is **damage to the tiny air sacs** in your lungs which has been **caused by tobacco smoke**. This damage causes these **air sacs to join together** and **trap pockets of air** inside the lungs. **Over time, small areas of the lung are destroyed**, so that instead of working lung, there are **empty holes**. **As more of the lung gets damaged**, this causes **difficulties with breathing**."*

### YOUR LUNGS SCRIPT (risk)

***"If you continue to smoke…***

*Some damage:* **…** If you continue to smoke, then this part of your lung **may become damaged in the same way**."

*No damage:* **…** While there is **no visible damage** to your lungs **yet**, if you continue to smoke, then this part of your lung is **likely to become damaged in the same way**."

### YOUR LUNGS SCRIPT (response efficacy)

***"If you stop smoking today…***

***Severe damage:*** … you will **immediately be protecting** these air sacs in your lungs and **keeping them working as well as possible**. This will help stop or slow down any further damage so that your breathing does not get worse…

***Some damage:*** **…** this part of your lung will **stay healthy**. You will **immediately be protecting** the healthy air sacs in your lungs and **keeping as many of them working** as possible…

***No damage:*** **…** this part of your lung will **stay healthy**…

***…It really is never too late to make positive changes.. After just three days, you should start to feel better in yourself."***

### YOUR LUNGS SCRIPT (picture 3)

***"This is a drawing of what a lung looks like inside…***

*The **darker parts** are the areas with **more damage** from smoking like in…the first picture of your lung/the second picture of the older smoker's lung…*

*The **lighter parts** are **healthy areas not damaged** by smoking/**areas with less damage** from smoking like in the middle/first picture of your lung."*

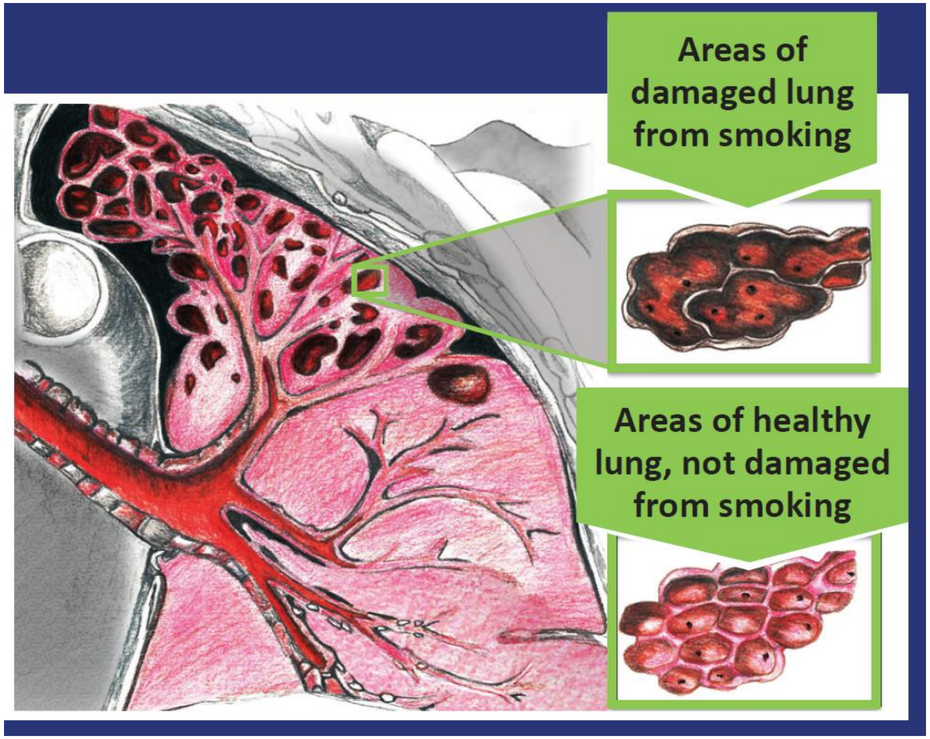

### INTERVENTION SCRIPTS:
### “Your heart”

### Example for 'some' damage

#### Your heart

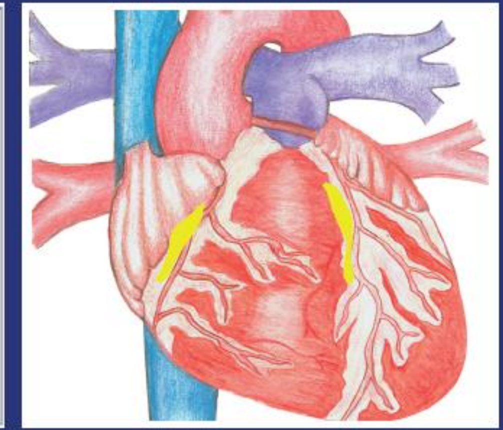

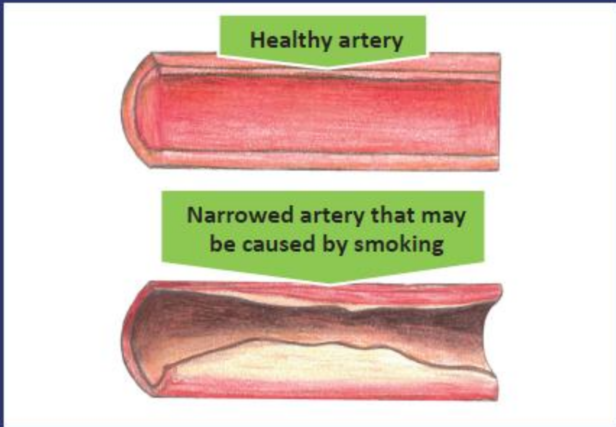


This is a picture of **your heart** taken from your CT scan.

The white areas show parts of the arteries going to your heart that have become hard or narrow.

Smoking is associated with the hardening and narrowing of the heart arteries. This can lead to problems including heart attacks.

This drawing shows what your heart looks like inside.

The yellow parts show roughly where the arteries going to your heart have become hard or narrow.

These are drawings of what healthy and narrowed arteries look like.

By stopping smoking, you can reduce the chance of the arteries to your heart becoming hardened or narrowed and lower your risks of problems like heart attacks.

### Presenting heart images

- Describe heart images
- Explain risk
- Clarify continued risk of smoking in context of result
  - Lay explanation of how coronary artery calcification is caused by smoking
  - Acknowledge not the only cause – to safeguard against scepticism which may undermine credibility (focus groups)

- Increase response efficacy
  - Immediate benefits of preventing/limiting smoking damage

- Manage potential for over-reassurance

### YOUR HEART SCRIPT (picture 1)

***"The first picture is a picture of <u>your own</u> heart…***

*Some damage:* …taken from **<u>your</u>** CT scan. The white parts of your scan picture are parts of the **<u>arteries going to your heart</u>** that have become narrow or hard, which is **<u>called "hardening of the arteries</u>**". "

*No damage:* ***…***taken from **<u>your</u>** CT scan."

### YOUR HEART SCRIPT (picture 2)

***"The second picture shows…***

*Some damage:* …**<u>what your own heart looks like inside</u>**. The white parts show roughly **<u>where the arteries going to your heart have become hard or narrow</u>.** You can see that there are **<u>also healthy parts of your arteries too</u>**, which haven't been damaged by your smoking **<u>yet</u>**."

*No damage:* ***…*** **<u>a heart that has been damaged by smoking</u>**. The white parts are where the arteries going to the heart have become hard or narrow."

### YOUR HEART SCRIPT (calcification)

- Lay explanation of coronary artery calcification describing **how** smoking damages the heart but acknowledging not the only factor

***"Hardening of the arteries means…***

…it is **harder for *your*/*the* blood to flow** through these arteries, which means that ***your*/*the*** **heart can't always get the supply of oxygen it needs to work properly** - increasing ***your*/*the*** chance of problems like **angina or heart attacks** in the future. There are **lots of factors** which may have caused this hardening and narrowing, but (***your***) smoking is one of the most important of them."

### YOUR HEART SCRIPT (picture 3)

***"These are drawings of healthy and narrowed arteries…***

By stopping smoking you can keep your arteries **as healthy as possible**, This is really important **at your age**.

*Some damage:* You can **reduce any further damage** from your smoking

*No damage:* You can **prevent any damage** from your smoking.  While there is **no visible damage** to your arteries **yet**, as a smoker you are at **high risk** of this damage in the future.

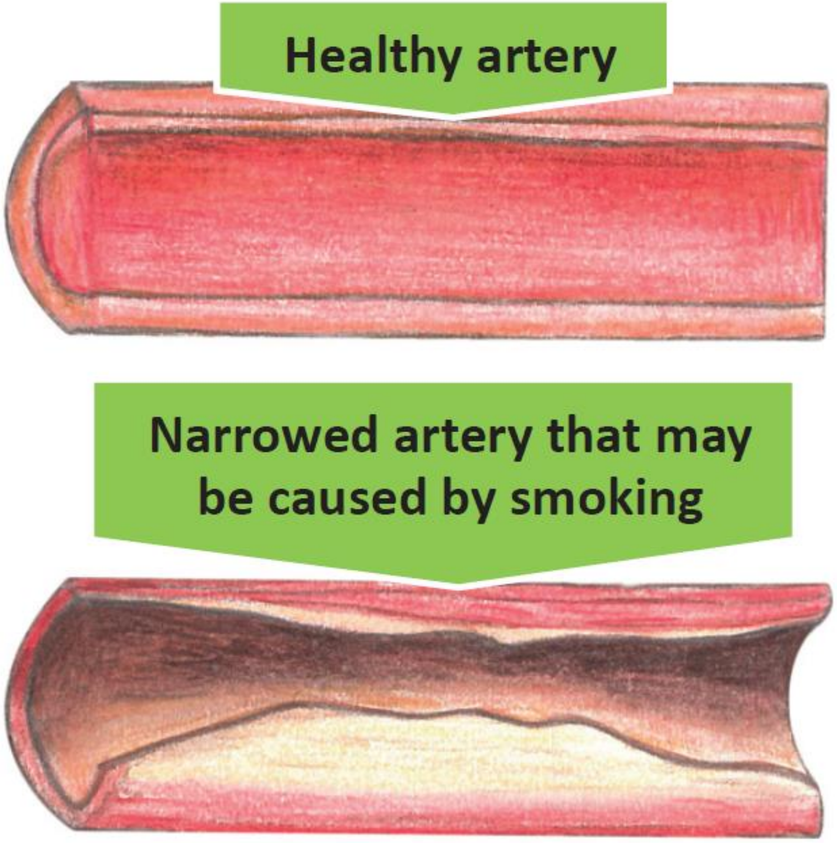

### YOUR HEART SCRIPT (picture 3)

***"Your heart will start to benefit from quitting very quickly…***

…In **just two weeks** from now, your **blood flow will start to improve**. This means a **healthier heart**, which is…

…***able to get a better supply of oxygen to help it work properly*** / ***not having to work as hard***…

…It also means you'll have a **lower chance** of problems like **heart attacks or angina**…

…In **just one year** from now, your **risk of heart disease** will go down to about half that of a smoker."

### INTERVENTION SCRIPTS:
### “How stopping smoking will help your health”

### Presenting immediate health benefits

- Increase response efficacy
  - Emphasise short-term benefits (especially for older age)
  - Less emphasis on longer-term benefits
  - Relate directly to the individual
  - Lay explanation for how health benefits accumulate to achieve significant reductions in risk

- Emphasise relevance for older age

### How stopping smoking will help your health

**After 20 minutes**
- Your heart rate goes back to normal.

**After 8 hours**
- Nicotine and carbon monoxide (a poisonous gas produced when smoking) in your blood goes down by half.
- Your oxygen levels go back to normal.

**After 2 days**
- You will not have any carbon monoxide left in your body.
- Your lungs will be clearer.
- You will be able to taste and smell better.

**After 3 days**
- You will find it easier to breathe.
- You will have more energy and walking will be easier.

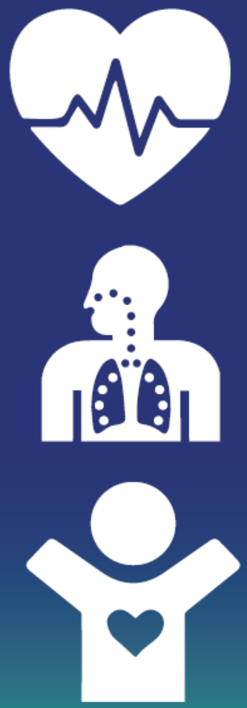

**After 2 to 12 weeks**
- Your blood will flow better around your body.
- This means more oxygen can get to important parts of your body.

**After 3 to 9 months**
- Your coughing and breathing will get better.
- Your lungs will start to work better- they can improve by up to 10%.

**After 1 year**
- Your risk of getting heart disease will go down to about half that of a person who is still smoking.

**After 10 years**
- Your risk of getting lung cancer will go down to about half that of a person who is still smoking.

**After 15 years**
- Your risk of having a heart attack will go down to the same as someone who has never smoked.

### HEALTH BENEFITS SCRIPT

“This page shows just **how much**, and **how quickly**, stopping smoking will improve your health, and **at any age no matter how long you have smoked**. The benefits start after just 20 minutes.”

Then talk through each health benefit

### HEALTH BENEFITS SCRIPT

***Explanations of nicotine and carbon monoxide***

"Nicotine is the **chemical** in tobacco that makes smoking **addictive**.

Carbon monoxide is a **poisonous gas** in cigarette smoke that **replaces the oxygen** in your blood stream. Breathing in carbon monoxide **can lead to heart disease**."

### HEALTH BENEFITS SCRIPT

***For those with no evidence of heart disease***

"**All of these positive changes** in your body mean that **a year from today**, your **risk of getting heart disease will go down to about half** that of a person who is still smoking**."**

### THANK YOU

**Questions:**
