## Additional File 3 for "Co-development of an evidence-based personalised smoking cessation intervention for use in a lung cancer screening context"

Appendix 3. GRIPP short reporting checklist. From: GRIPP2 reporting checklists: tools to improve reporting of patient and public involvement in research

| Section and topic | Item | Reported on page No |
|---|---|---|
| 1: Aim | Report the aim of PPI in the study | 4-6 |
| 2: Methods | Provide a clear description of the methods used for PPI in the study | 8-9 |
| 3: Study results | Outcomes—Report the results of PPI in the study, including both positive and negative outcomes | 9-12 |
| 4: Discussion and conclusions | Outcomes—Comment on the extent to which PPI influenced the study overall. Describe positive and negative effects | 13-16 |
| 5: Reflections/critical perspective | Comment critically on the study, reflecting on the things that went well and those that did not, so others can learn from this experience | 15-16 |
